# Global Transportability of Clinical Trial Outcomes to Real-World Lung Cancer Populations A case Study using Lung-MAP S1400I

**DOI:** 10.1101/2025.05.30.25328679

**Authors:** Alind Gupta, Nicholas Latimer, Manuel Gomes, Kelvin Chan, Seamus Kent, Stephen Duffield, Sreeram Ramagopalan, Eran Bendavid, Winson Cheung, Vivek Subbiah, Paul Arora

## Abstract

**Importance:** The relevance of randomized clinical trials (RCTs) outcomes to real-world settings – especially across countries – is sometimes limited by their restrictive eligibility criteria and variations in standards of care compared to routine clinical practice.

**Objective:** To assess the transportability of findings from the RCT Lung-MAP S1400I to real-world populations in the United States (US), Germany and France.

**Design:** This empirical validation study used patient-level data from the RCT Lung-MAP S1400I to build a transportability model to adjust for differences in patient characteristics from real-world target patient populations. Two sets of adjustments were performed – one limited to the set of measured clinical variables, and the second additionally including external information drawn from published literature and substantive knowledge on patient subgroups excluded from the trial. The latter enabled transportability to a significantly more diverse and representative real-world patient population by relaxing the stringent exclusion criteria used in Lung-MAP S1400I. For benchmarking, we compared how well the transportability analysis approximated observed overall survival in the respective real-world cohorts.

**Setting:** Observational study.

**Participants:** Eligible individuals diagnosed with advanced or metastatic NSCLC and previously treated with systemic therapy.

**Intervention/exposure:** Nivolumab monotherapy.

**Main outcome measures:** Overall survival.

**Results:** Sample size for the nivolumab arm in Lung-MAP S1400I was 127 and ranged from 133 to 1051 for the various real-world cohorts included. Patients with ECOG scores of 2+, index cancer stage <IV, presence of *ALK*/*EGFR* mutations, presence of comorbidities and prior exposure to immunotherapy/targeted therapies were excluded from Lung-MAP S1400I, were but were eligible to receive nivolumab monotherapy in real-world care. Adjusting for measured clinical differences improved alignment of patient outcomes in the RCT and the real-world cohorts. However, only when variables related to excluded patient groups were also addressed did the results fully satisfy control conditions, yielding the closest approximation to real-world survival in the US, Germany, and France (mean discrepancy: 0.27 months over ∼30 months).

**Conclusions:** Overall survival in a more diverse real-world patient population could be extrapolated using data from the Lung-MAP S1400I trial when complemented with external information about excluded patient groups.

## Introduction

Although randomized clinical trials (RCTs) are key to understanding questions about the efficacy of novel treatments, their generalizability to routine clinical care is sometimes thought to be limited (Rothwell 2005; Kennedy-Martin et al. 2015). By generalizability, we refer to the extent to which the patients and outcomes from an RCT approximate those typically observed in routine clinical practice (Schwartz and Lellouch, 1967). Although regulatory and reimbursement decisions are driven primarily by evidence from RCTs, there is a growing interest in understanding the generalizability of RCTs to contextualize their results for decision-making, and to optimize equity and value in healthcare and patient outcomes (Inoue & Hsu 2024; de Jonghe et al. 2014; see also draft guidance for diversity action plans from the United States Food & Drugs Administration). Similarly, there is increasing interest in the use of real-world data (RWD) to inform regulatory and reimbursement decisions, and there has been a growing use of external control arms, where RWD is used to emulate a control arm for a clinical trial (Mishra-Kalyani et al. 2022). When either RCT or RWD evidence under consideration is from a different jurisdiction or country, issues around generalizability become even more important.

Several factors can impact the generalizability of RCTs, including the use of restrictive eligibility criteria, differences in standards of care from routine care, and adherence to treatment. RCTs often select healthier patients (e.g., Rogers et al. 2021; Anderson et al. 2020) who may be more likely to adhere to the study’s protocol, and to mitigate safety concerns associated with treating patients with poor performance or multiple comorbidities. These factors can limit an understanding of whether RCT results can be extrapolated to those patients typically seen in clinical practice. Regardless of the exclusion criteria, if the treatment effect differs according to patient/treatment characteristics (e.g., by age or subsequent therapy), then the treatment outcomes in the RCT may not be an accurate measure of real-world patient outcomes if the distributions of patient characteristics in the RCT is different from that in the real-world. A discussion of generalizability in the presence of effect heterogeneity, non-collapsibility of effect measures and the use of conditional versus marginal estimands is out of scope for this paper but can be found elsewhere (e.g., Remiro-Azócar 2024).

A fundamental barrier to quantifying RCT generalizability is that real-world patients who receive those therapies are often more diverse than patients enrolled in RCTs – i.e. the RCT may exclude certain patient subgroups. For example, patients with cancer who have an ECOG performance scores of 3 or 4 (considered frail) are often excluded from oncology trials but may sometimes receive systemic cancer therapy in the real-world (e.g., see real-world studies described later in this paper). How can we estimate outcomes for these patients not included in RCTs, i.e., “transport” results from the trial to these patients (we refer to this as “transportability” in this manuscript)? In causal inference terminology, this issue is called lack of “positivity” because the probability of observing excluded real-world patient groups in the RCT is, by definition, zero. Consider an RCT composed of patients from the United States that was submitted as evidence of treatment effectiveness to a Japanese decision-making body. There may be few to no Japanese patients in the RCT, and therefore it is not straightforward to use them to understand how Japanese patients will respond to treatment. Often, an assumption is made that the variables or variable categories underlying lack of positivity are unrelated to the outcome, or do not modify the treatment effect measure, after which the problematic variable can be ignored. However, this assumption is often untenable; it is known, for example, that Asian patients respond differently to some targeted therapies for non-small cell lung cancer compared to non-Asian patients, partly due to differences in *EGFR* variants (Kim et al. 2021; Zhou et al. 2011), and therefore racial differences underlying lack of positivity cannot be ignored in this example.

The generalizability of RCTs has been a long-standing subject of interest – Rothwell (2005) documented barriers to generalizability, such as selective recruitment, unrepresentative trial contexts, and narrow outcome definitions. The empirical evaluation of the generalizability of RCTs is a more recent area of research, fueled partly by the popularization of causal inference methods and interest in real-world evidence (Stuart et al. 2015; Degtiar and Rose 2021). Existing empirical studies of RCT findings have evaluated the generalizability of relative effect estimates only; in contrast, absolute measures of risk are often more useful in the context of ECA analyses or indirect comparisons. Furthermore, there is often no systematic assessment of causal assumptions or missing data, and individual-level are typically used (see Levy et al. 2024 for a systematic review of existing transportability studies). Importantly, the lack of positivity, which is likely to affect many assessments of the real-world applicability of RCTs, has not been assessed in practice so far. External information about potential unmeasured variables is also often not considered In this benchmarking study, we attempted to quantify the extent to which results from a US-based RCT generalize to real-world patient populations across the world, after accounting for differences in risk factors between them. We used publicly accessible data from the phase III RCT Lung-MAP S1400I (Gettinger et al. 2021), which included patients with stage IV squamous cell lung cancer who had previously received treatment. It compared the efficacy of nivolumab, a popular anti-PD-L1 immune checkpoint inhibitor, as monotherapy versus in combination with ipilimumab, a CTLA-4 inhibitor. Based on an intention-to-treat analysis, the trial found that the combination therapy did not significantly improve overall survival compared to nivolumab alone (hazard ratio 0.87, 95% confidence intervals [0.66, 1.16]). Because the use of this combination is not authorized for this indication, it is not possible to directly benchmark the comparative treatment effect. Instead, we evaluated the transportability of overall survival curves on nivolumab in Lung-MAP S1400I to real-world patients eligible to receive nivolumab in the United States, France and Germany using published data on outcomes from real-world studies.

Our study offers three main methodological contributions to the field. First, we propose a transparent study design specification for transportability analyses based on the target trial framework. The target trial framework is a design approach that is typically used for emulating a hypothetical randomized trial to improve the clarity, transparency, and validity of observational analyses (Hernán et al. 2022); here, we use it to explicitly document the alignment of eligibility criteria, treatment regimens and follow-up across different studies in the context of transportability. Second, our study implements synthesis estimators for handling positivity violations (Zivich et al. 2024) in a real-world application.

Similar approaches can also incorporate external information to inform unmeasured variables (Gupta et al. 2025) or missing values when using summary-level data; in practice, these issues are often ignored or circumvented by altering variable definitions or eligibility criteria instead. Third, we use several controls. Controls refer to pre-specified scenarios – either positive controls where transportability is expected, or negative controls where it is not – that can be used to benchmark and contextualize the main findings (also see *Appendix – Statistical formalism: Controls*). These can help assess internal consistency and provide a reference for interpreting results across different real-world data sources within the same country.

## Methods

### Trial data

Lung-MAP S1400I (NCT02785952; Gettinger et al. 2021) was a US-based open-label randomized phase III sub-study within the Lung-MAP (SWOG S1400) master protocol conducted between 2015 and 2018. The trial compared nivolumab monotherapy versus a nivolumab/ipilimumab combination in 127 and 125 patients, respectively, recruited in the United States. Eligible patients had metastatic NSCLC with squamous cell histology whose tumors lacked actionable genomic alterations and who had previously progressed on platinum-based chemotherapy (Gettinger et al. 2021). The primary endpoint was overall survival and secondary endpoints included progression-free survival and response. Over a follow-up of approximately 36 months, the hazard ratio for all-cause mortality comparing the combination therapy to nivolumab alone was found to be 0.87 [0.66, 1.16].

For this study, a pseudonymized patient-level dataset for 252 randomized patients from Lung-MAP S1400I was accessed through Project Data Sphere courtesy of the National Cancer Institute (NCI). We did not have access to the complete trial dataset in a standard format – the available dataset contained what we assumed were a subset of the complete set of variables originally measured in the trial.

Justification for the choice of Lung-MAP S1400I for this study can be found in the study protocol available online at https://www.medrxiv.org/content/10.1101/2024.05.25.24307916v2.

### Real-world data sources

We identified published real-world studies of patients with NSCLC treated with nivolumab in the second-line setting in the United States, Europe and Asia through a targeted literature search. A description of the seven studies identified from the United States (Stenehjem et al. 2021), Germany (Martin et al. 2022; Chouaid et al. 2022), France (Chouaid et al. 2022; Barlesi et al. 2020), England (Snee et al. 2021) and Japan (Morita et al. 2020) can be found in the *Appendix – Study specifications: Data provenance*.

Note that we identified additional real-world studies of nivolumab from Japan, South Korea, Netherlands and France during the literature search. However, because these studies did not report crucial elements, such as baseline variables and Kaplan-Meier curves for overall survival stratified by histology, we did not include them.

### Eligibility criteria

Because this study uses published information from analyses of real-world patient cohorts, eligibility criteria for the target real-world populations were essentially predetermined and unalterable. The target real-world populations of interest in this study therefore are defined as patients with squamous-cell NSCLC who are eligible to receive nivolumab in routine clinical care in their respective countries and who meet any additional clinical inclusion/exclusion criteria that were applied by the respective real-world studies. A formal comparison of eligibility criteria between Lung-MAP S1400I and the seven real-world studies included in this study can be found in the *Appendix – Study specifications: Target trial specification*. The target trial specification table also highlights both known (i.e., explicitly reported) and potential (i.e., not explicitly reported, thus assumed) structural positivity violations where the eligibility criteria applied in Lung-MAP S1400I were more restrictive than those in real-world studies.

### Variables

The choice of variables for adjustment was based on a causal diagram representing our assumptions about the setting and clinical expertise in the team (*Appendix – Causal graph*). A comprehensive description of variables, including their categorizations, as well as their values, and comments about harmonization and measurement, is provided in *Appendix – Study specification: Variables*. The same table also includes summary-level values for unmeasured variables based on external information; the source and justification for these values is provided.

### Controls and Sanity checking

In this study, we use the term “controls” to refer to pre-specified scenarios selected to test the expected presence (positive controls) or absence (negative controls) of transportability. By design, positive controls are settings where transportability should hold, while negative controls are settings where it should fail. These controls serve to benchmark the main findings, assess their internal consistency, and provide context for interpreting results across multiple real-world data sources from the same country. For details, please see *Appendix – Statistical formalism*.

As a positive control for transportability analysis, we use the Lung-MAP S1400I trial itself as the target population, where we expect excellent alignment between our predictions (i.e., model-based estimates) and actual survival (see for example Ramagopalan et al. 2022). As described in the statistical formalism, identifiability conditions are trivially satisfied when transporting from this trial to itself. We expect that a model fitted on Lung-MAP S1400I as the “source” study population should be able to estimate survival for the same Lung-MAP S1400I cohort very well with a well-specified model. Therefore, this control ensures that our modelling approach is technically valid.

As negative controls, we use cohorts from real-world studies in England (Snee et al. 2021) and Japan (Morita et al. 2020) (see caveat about the English population as a control in *Appendix – Statistical formalism*). England and Japan will henceforth be shown with an asterisk (*) as “England (*)” and “Japan (*)” to mark them as negative controls. For the English study, patient characteristics and overall survival are reported from the time of cancer diagnosis, not nivolumab initiation. Patients in the Lung-MAP study are the select set of resilient patients who have not only survived until second-line but are also still fit to receive systemic therapy at that point, and therefore as a population, should have much better prognosis than a patient cohort at the time of initial diagnosis of which 62.5% were fit enough to receive systemic therapy (Snee et al. 2021), above and beyond the factors we are adjusting for in this study to account for differences between countries at the time of start of second-line treatment. Therefore, we expect that our model-based survival estimates for England (*) should be better than (i.e., overestimate) the survival reported in the study from England. Note that for technical reasons described in *Appendix – Statistical formalism*, results for England* are presented only in supplementary tables and figures.

On the other hand, for Japan (*), survival is reported only for the combined NSCLC population, including squamous, non-squamous and other histologies. Non-squamous histology accounts for the majority (∼80%) and is associated with better prognosis than squamous histology in NSCLC patients. Our study plan is not sufficient to transport from squamous cell histology (Lung-MAP S1400I only includes squamous cell histology) to all NSCLC. Therefore, we expect that our model-based survival estimates for Japan (*) should be worse than (i.e., underestimate) the survival reported in the study from Japan.

We also included a real-world cohort from the United States (Stenehjem et al. 2021) with the hypothesis that if our approach works, then we should get good generalizability at least from a US-based RCT to the US real-world population. Note that this was not a positive control per se; we only expected that, comparatively, there should be better transportability to the US real-world population than to real-world populations in Europe. Note that the US study reported Kaplan-Meier curves for squamous NSCLC stratified by ECOG performance scores, rather than an overall curve. Therefore, for benchmarking based on restricted mean survival time, we selected the curve corresponding to ECOG scores 0-1. However, the study did report the overall median survival for the squamous histology subgroup, which we used for benchmarking based on median overall survival. Finally, we also included two real-world cohorts each from Germany and France to assess whether transportability results are consistent between different databases within the same country. This helped us evaluate the internal consistency of our findings and the impact of data provenance or measurement across databases on the results.

### Multiple imputation and handling missing values

In the NCI dataset for Lung-MAP S1400I, 36.1% of patients (n=91 out of 252) contained missing values for PD-L1 expression. Our exploratory analyses with the positive control indicated that complete-case analysis yielded relatively anomalous parameter estimates for the G-formula model used for the substantive analysis (the G-formula is described below), and single imputation of PD-L1 was not sufficiently reliable at capturing uncertainty in resulting estimates (see *Appendix – Sanity checking*).

Multiple imputation was implemented in R using the *mice* package (van Buuren et al. 2011), which uses chained equations for a fully conditional specification of the joint distribution of variables. All variables used in the substantive analysis, including the treatment and outcome variables, were included in the multiple imputation model for PD-L1, which was modelled using logistic regression. Based on results from exploratory analyses, 10 imputations were sufficient for recapitulating mean survival.

For real-world studies, completely random missingness was assumed because only summary-level data on marginal distributions of variables, including missing values, was available, and therefore imputation was not feasible. For example, if for a total of 100 patients for which overall survival was reported, 50 patients (50%) were reported to have an ECOG performance score of 0-1, 20 (20%) had an ECOG score of 2 or higher and the value was missing for the remaining 30 patients (30%), then the total prevalence for ECOG 0-1 would be n=50 patients plus 50% of 30 (n=15), equalling 65% (i.e., 50 + 15 = 65 divided by 100 patients). That is, we assumed that the distribution was identical in patients with missing values (see *Discussion – Limitations*).

### Copula simulation

Because only summary-level covariate data from the real-world populations was available, a copula was used to model the joint distribution of covariates and to sample synthetic data with marginals matching those observed. This synthetic patient-level data can then be used for covariate adjustment in the substantive analysis, with multiple replicates to account for the uncertainty in sampling. The process for copula simulation is described below.

A multivariable Gaussian copula was fitted to baseline patient covariates from Lung-MAP S1400I data. Justification for copula and copula parameter choice is provided in *Appendix – Sanity checking*. The fitted copula was then used to simulate patient-level covariate data from target populations based on their corresponding summary statistics, i.e., the (population-level) mean and standard deviation for continuous variables, which were assumed to be Normally distributed, and prevalence for dichotomous variables, which were modelled using the binomial distribution. In essence, copula simulation produced synthetic patient-level datasets that matched the marginal distributions of covariates in the target populations but simulated the correlations between those covariates based on the covariance structure of the Lung-MAP data (see sensitivity analyses in *Appendix – Sanity checking: Copula choice*). The *copula* package in R was used for the copula analysis. The copula approach has previously been used for population adjustment in indirect treatment comparisons for a similar purpose (Phillippo et al. 2020).

### Parametric G-formula

The g-formula is a method for estimating marginal outcome distributions under interventions by adjusting for measured covariates. In this setting, the g-formula can be used to adjust for differences in covariate distributions between populations by standardizing a model fitted to data from the source population to the distribution of covariates in the target population (Rose & Degtiar 2022). We used the g-formula here because it can offer better statistical efficiency (at the risk of bias due to misspecification) in settings with limited covariate overlap between study and target populations, whereas inverse weighting would result in very low effective sample sizes given our already modest sample size (data not shown).

We used a prespecified outcome model (“Q-model”) for overall survival as a function of patient-level covariates. A pooled logistic regression model was used to model survival. Assuming a sufficiently rare monthly incidence rate for outcome (i.e., all-cause death) in the population, the parameters of the pooled logistic regression model approximate those from a Cox proportional hazard model (see *Appendix – Sanity checking: Equivalence between G-formula and Cox proportional hazards coefficients*). To reduce risk of model misspecification from an overly parsimonious Q-model, we included squared terms for continuous variables, and 2-way interactions between all covariates in the Q-model. Time was modelled using polynomial terms up to a degree of 4. The parameters of the Q-model were estimated from Lung-MAP S1400I up to a maximum of 35 months of follow-up to produce our fitted “transportability model”. This transportability model was used to estimate survival probabilities standardized to the covariate distributions in the target population (see also *Appendix – Justification for other study design choices: Selection of patient cohorts representing the target population*) to obtain marginal, i.e., population-level, survival probabilities. To estimate 95% confidence intervals, 500 replicates of non-parametric bootstrapping were implemented (see *Appendix – Sanity checking: Number of iterations* for justification). Hazard ratios over the follow-up could then be calculated based on the estimated standardized survival on nivolumab versus nivolumab+ipilimumab combination.

Three sets of analysis were performed, which as shorthand we refer using the terms CRUDE, BASE and POS. The CRUDE results present raw unadjusted results for survival with no statistical adjustment. BASE represents a base case analysis where we adjust only for measured variables. Mismeasured variables were excluded. For POS, we account for well-measured variables as in BASE, but also any positivity violations, including corrected mismeasured variables. A description of synthesis estimators for addressing positivity violations can be found in Zivich et al. (2024) and in the study protocol. Briefly, information for variables underlying positivity violations, i.e., for variables that were excluded from Lung-MAP S1400I but not in the real-world studies, such as ECOG performance scores of 2 or higher, the synthesis model included the BASE transportability model with additional parameters that needed to be specified based on external information, such as published multivariable analyses. In the case of ECOG performance score of 2 or higher, for example, this coefficient corresponds to the (log) hazard ratio for all-cause mortality associated with having an ECOG score of 2 or higher versus an ECOG score of 0-1, the latter representing the inclusion criterion in Lung-MAP S1400I. For the additional terms included in the POS analysis, we included only main effects. The complete list of these variables, their coefficients for the POS model, associated references and our rationale for their values are described in *Appendix – Study specifications: Synthesis parameters.* Published studies which contained quantitative information on prevalence and/or prognostic association with mortality that could inform the POS model were identified using targeted search terms – a systematic review was not performed.

### Benchmarking

Benchmarking could only be performed for overall survival on nivolumab. We overlaid plots of transportability model-based estimates in the target real-world populations with the actual observed survival in the corresponding real-world cohorts. Overlap between these two was assessed qualitatively via visual inspection of the curves. As a quantification of the differences, we computed mean survival up to the maximum observed follow-up (“restricted mean survival time” or RMST), which is equivalent to the area under the survival curves, and then calculated the difference between the mean survival between our model-based estimates and the observed real-world survival for each target population. Mean difference in RMST and 95% confidence intervals were documented. Median overall survival estimates were also compared.

## Results

### Patient characteristics

Patients in Lung-MAP S1400I were, on average, younger than those in real-world cohorts (Table 1). As expected, clinical characteristics were more similar between Lung-MAP S1400I and the US real-world cohort than to patient cohorts in Germany or France.

**Table 1.** List of measured baseline characteristics. The standardized mean difference (SMD) is the average of pairwise SMDs compared to the trial, excluding any missing values (-).

| Risk factor | Lung-MAP<br>S14001 | USA<br>(Flatiron) | Germany<br>(ENLARGE<br>-Lung) | Germany<br>(CRISP) | France<br>(ESME-<br>AMLC) | France<br>(EVIDENS) | Japan<br>(*) | England<br>(*) | Average<br>SMD |
| --- | --- | --- | --- | --- | --- | --- | --- | --- | --- |
| Sample size | 127 | 1051 | 242 | 567 | 133 | 437 | 901 | 819 | N/A |
| Age in years,<br>mean (SD)<sup>†</sup> | 66.8 (8.53) | 69 (8.7) | 70 (15) | 69 (8.9) | 66 (9.5) | 68 (11.75) | 67 (15) | 70.8 (9.4) | 0.21 |
| Sex = male (%) | 67.1 | 63 | 71.8 | 70.7 | 82 | 80.8 | 72.3 | 61.7 | 0.17 |
| Race = white (%) | 81.7 | 79 | - | - | - | - | - | - | 0.07 |
| ECOG score = 0<br>or 1 (%) | 100 | 70.1 | 86.1 | 98.1 | 95.5 | 79.2 | 81.3 | 51.7 | 0.68 |
| Cancer stage at<br>diagnosis = IIIB+<br>(%) | - | 88.6 | 87.8 | 92.9 | 77.9 | 96.6 | 66.5 | 48.3 | - |
| Smoking status =<br>ever (%) | 98.8 | 96.3 | 88.8 | 94.2 | 98 | 94.7 | 79.9 | - | 0.3 |
| Bone metastases =<br>yes (%) | - | - | 22.8 | 37.6 | 30 | - | 28.4 | - | - |
| Liver metastases =<br>yes (%) | 21 | - | 14.1 | 21.8 | 20.6 | 15.3 | 11.5 | - | 0.12 |
| Brain metastases =<br>yes (%) | 7.94 | 6 | 11.6 | 12 | 19.6 | 10.3 | 22.3 | - | 0.2 |
| PD-L1 expression<br>= >1% (%) | 60.9 | 51.3 | 64.6 | 38.1 | 43 | 61.6 | 52.9 | - | 0.21 |
| EGFR mutation =<br>yes (%) | 0 | 1.72 | 18.3 | 1.5 | 0.4 | - | 15.3 | - | 0.34 |
| ALK<br>rearrangement =<br>yes (%) | 0 | 0 | - | 0.8 | 0.5 | - | 1.8 | - | 0.14 |
| Index line =<br>second line (%) | 83.7 | 85 | 81.9 | 91.7 | 75.7 | 73.8 | 51 | N/A | 0.25 |

<sup>†</sup> Mean and standard deviation were approximated from median, range or interquartile ranges using standard approximations if only the latter were reported.

Patient characteristics across different real-world cohorts in Germany and France were not wholly consistent even though the studies captured patient data spanning similar time periods (roughly 2016-2019; see *Appendix – Study specification: Data provenance*). For example, despite similarities in patient age distributions, the ENLARGE-Lung and CRISP cohorts from Germany differed with respect to the percentage of PD-L1-positive patients (64.6% versus 38.1% respectively in Table 1, based on a completely-at-random assumption for missing data). Similarly, the two French studies also reported different proportions for PD-L1 positive patients (43% and 61.6%, Table 1). They also differed in the distribution of ECOG performance scores at baseline and in the proportion of patients diagnosed with early-versus late-stage cancer (Table 1). The reported proportion of patients with *EGFR* mutations in ENLARGE-Lung was also uncharacteristically high for squamous cell NSCLC (18.3%); this was likely due to mismeasurement (see Sebastian et al. (2022) for their discussion on *EGFR* testing in their study).

The French ESME-AMLC cohort was unusual in that it contained a relatively high number of patients diagnosed with early-stage (stage <IIIB) NSCLC (22.1% versus an average of 8.5% in the rest of non-controls); notably, among the French and German studies, ESME-AMLC was the only study conducted retrospectively using an existing database and it also had the smallest sample size (n=131).

As for the controls, the Japanese (*) cohort (n=901), which included patients with both squamous and non-squamous NSCLC, had fewer smokers (80%) and a higher prevalence of *EGFR* mutations (15.3%) compared to non-controls; this was expected due to the contribution of the non-squamous histology. The English (*) study (n=819) did not report many baseline patient characteristics, but it had a higher proportion of patients with favorable ECOG performance scores, which was also expected given that it captured patient characteristics at the time of cancer diagnosis rather than at second line like the rest.

Structural positivity violations resulted from the presence of patients with ECOG scores of 2+, index stage <IV, presence of *ALK*/*EGFR* mutations, presence of comorbidities and prior exposure to immunotherapy or targeted therapies in all real-world initiators of nivolumab; these were all exclusion criteria in Lung-MAP S1400I (*Appendix – Study specification: Target trial specification*). Identifiability assumptions are described in *Appendix – Identifiability conditions*.

### Comparison of crude survival

As shown in Figure 1, the unadjusted (“CRUDE”) survival in Lung-MAP S1400I was on par or better than overall survival in the non-control cohorts from the United States, Germany and France. This was consistent with the fact that patients with performance scores of 2 or higher were excluded from Lung-MAP S1400I but were present in all real-world cohorts. Interestingly, however, by month 30, crude survival in the trial was in fact lower than in the US, Germany and France. This may partly be a result of the small at-risk sample in the trial by that time (n=9 in the nivolumab arm) leading to imprecise model estimates. Overall survival in the control cohort behaved expectedly – the Japanese (*) cohort showed better survival than Lung-MAP S1400I and the non-controls (Figure 1) English (*) cohort showed significantly worse survival than the rest. The restricted mean survival times estimated from crude Kaplan-Meier curves are shown in Table 2 and median overall survival in Supplementary table 1.

**Figure 1.**
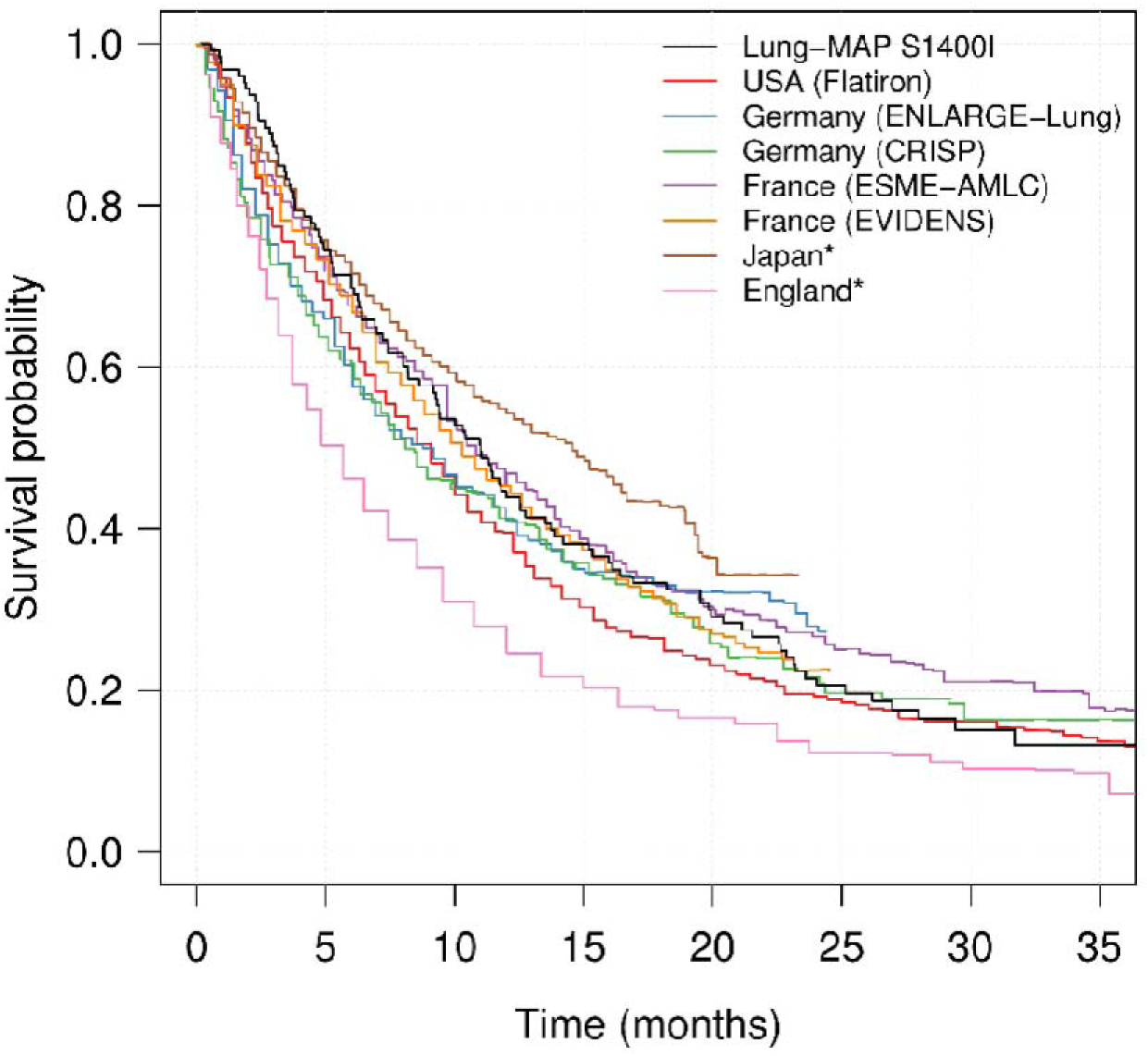
Crude survival in Lung-MAP S1400I and published real-world cohorts.

**Table 2.** Restricted mean survival time (RMST) over the follow-up shown as a difference between predicted and observed overall survival. The average was calculated for populations in USA, Germany and France (5 cohorts in grey) only, excluding control cohorts.

| Target population | Follow-up time (months) | Difference in predicted RMST versus actual RMST (months) |  |  |
| --- | --- | --- | --- | --- |
|  |  | CRUDE | BASE | POS |
| Lung-MAP S14001 | 35 | 0 | 0.40 [-2.50, 3.02] | 0.56 [-1.71, 3.02] |
| USA (Flatiron) | 35 | -1.63 | 1.87 [-0.80, 3.91] | 0.46 [-2.15, 2.79] |
| Germany (ENLARGE-Lung) | 23 | -1.01 | 0.24 [-1.56, 2.34] | -0.03 [-2.22, 2.33] |
| Germany (CRISP) | 35 | -1.27 | 0.61 [-1.61, 4.27] | 0.52 [-2.43, 3.61] |
| France (ESME-AMLC) | 35 | 0.62 | -1.30 [-3.84, 1.35] | -2.09 [-4.03, 1.45] |
| France (EVIDENS) | 23 | -0.59 | 0.37 [-1.39, 1.98] | -0.29 [-2.13, 1.35] |
| Japan* | 22 | 1.13 | -1.77 [-4.10, 0.51] | -1.61 [-5.46, 1.25] |
| **Average across USA, Germany and France:** | 30.2 | -0.78 [-1.54, -0.01] | 0.39 [-0.58, 1.36] | -0.27 [-1.30, 0.77] |

A difference in RMST of zero implies that model-based survival estimates were identical to observed survival estimates in the target population on average. A negative RMST difference implies that the observed survival in target populations was better on average than we were able to estimate using the transportability model, and vice versa for a positive RMST difference.

As described in the previous section, patients in Lung-MAP S1400I and in the seven real-world cohorts differed in the distribution of several risk factors for all-cause mortality. Therefore, a crude comparison of the trial and real-world overall survival estimates to assess the generalizability of Lung-MAP S1400I could result in biased conclusions. We next attempted to adjust for these differences using transportability analysis.

### Transportability model results

We first adjusted for measured variables only (“BASE” analysis). This is the standard analysis that is often used in practice. Results for adjusted survival curves from the BASE analysis are shown in Figure 2. We found that adjustment for measured variables improved the approximation of real-world survival estimates relative to the CRUDE case with no adjustment. The corresponding restricted mean survival is quantified in Table 2, and it corroborated the improvement (mean survival discrepancy was - 0.78 months for CRUDE and 0.39 months for BASE across non-controls, with a value of 0 being ideal). The median survival estimates are shown in Supplementary table 1.

**Figure 2.**
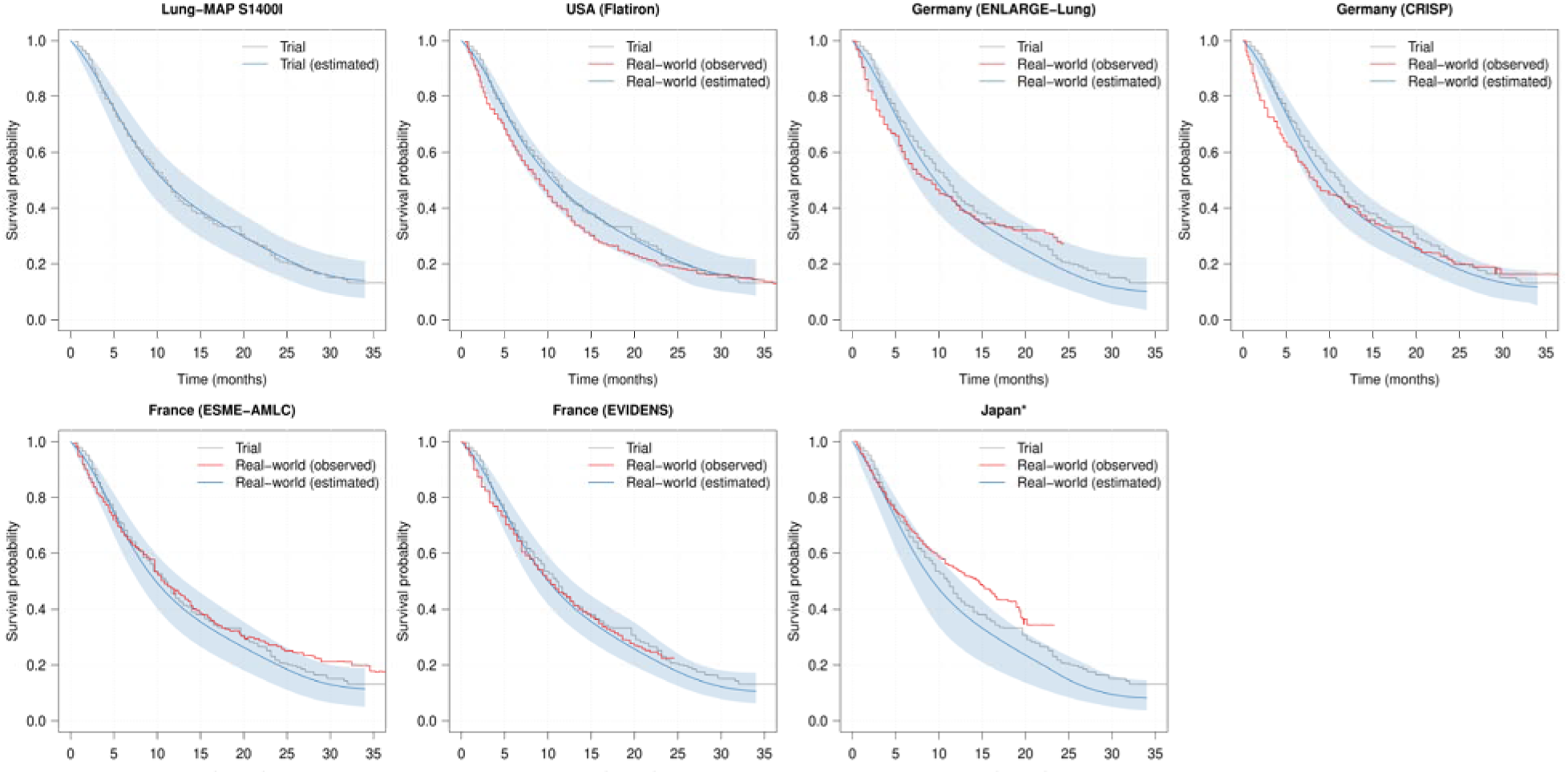
Results from the BASE analysis after adjusting for measured variables only. The grey curve represents crude survival in the Lung-MAP trial, which is the same across all panels. Shaded blue regions represent 95% confidence intervals. Model-based estimates (blue) are plotted along with the observed survival in the real-world cohorts (red).

Both the positive and negative controls behaved expectedly – model-based survival estimates almost perfectly approximated survival in Lung-MAP S14001 and significantly underestimated survival in Japan (*) (Figure 2). Nonetheless, the BASE model approximated survival better in France and Germany compared to the United States (Figure 2 and Table 2). This was counterintuitive because it is more likely that a US trial generalizes better to the US real-world population, particularly given the similarity of reported baseline clinical characteristics, than to real-world populations in other countries. This finding suggested that adjustment for measured variables alone may not be sufficient for a good transportability model.

Finally, we additionally accounted for mismeasured and unmeasured variables, and variables underlying structural positivity issues in the “POS” analysis – these variables were ECOG PS, smoking status, presence of EGFR/ALK mutations, cancer stage at index date, presence of comorbidities and prior exposure to immuno-or targeted therapy. The positive and negative controls behaved expectedly, that is, in line with our hypotheses (Figure 3). Additionally, we found a near-perfect alignment to survival in the US real-world population in the POS analysis (Figure 3). Quantification using mean survival showed that the POS analysis was able to approximate the mean survival better than the BASE model for the German and French cohorts except ESME-AMLC (Table 2 and Supplementary table 1). We also found internal consistency in results across different datasets within the same country – the POS transportability model slightly overestimated real-world survival in both German cohorts early during follow-up, but underestimated survival in both French cohorts (Figure 3). Based on these results, the POS model was preferable to the BASE model. Hazard ratios comparing nivolumab versus nivolumab/ipilimumab combination are provided in Supplementary table 2 with broad consistency with findings from Lung-MAP S1400I suggesting that effect modification on the hazard ratio scale was not substantial; interestingly, the effect size was largest (farthest from the null) in the French populations.

**Figure 3.**
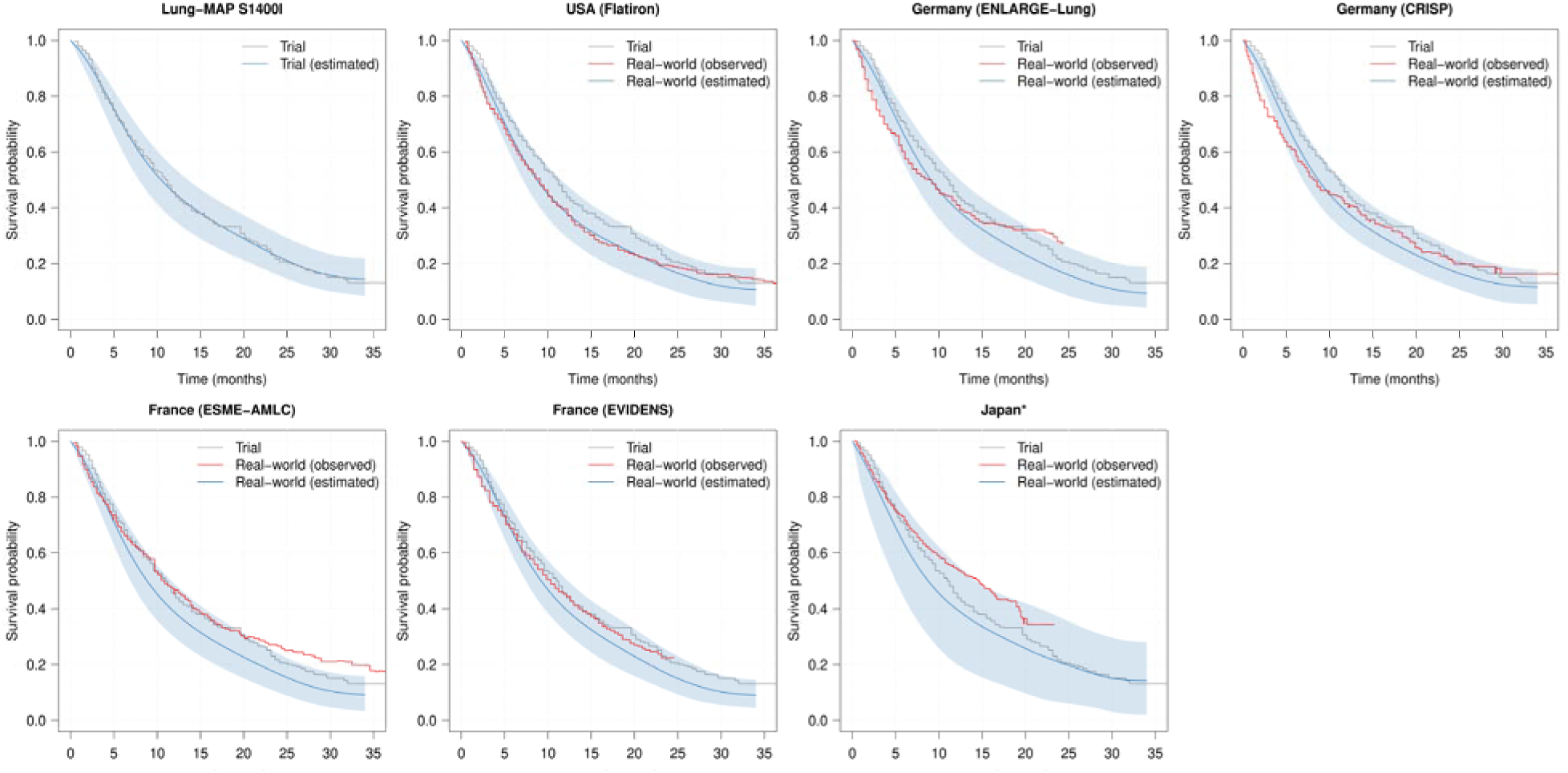
Results from the POS analysis after accounting for unmeasured variables and positivity violations. The grey curve represents crude survival in the Lung-MAP trial, which is the same across all panels. Shaded blue regions represent 95% confidence intervals. Model-based estimates (blue) are plotted along with the observed survival in the real-world cohorts (red); a high degree of overlap between the blue and red curves reflects high transportability.

## Discussion

Here, we present findings from a case study evaluating the transportability of patient outcomes from the Lung-MAP S1400I trial to real-world populations in the United States, Germany and France. Lung-MAP S1400I enrolled patients with metastatic squamous cell NSCLC – a relatively understudied histological subtype with distinct characteristics and poorer prognosis than the more prevalent non-squamous forms. As is common in clinical trials in oncology, Lung-MAP S1400I employed eligibility criteria that restricted enrollment to a subset of real-world patients with squamous NSCLC, presumably those for which the therapies were assumed to be most efficacious and safe. We found that the trial and real-world cohorts of patients eligible to receive nivolumab monotherapy for this indication differed in the distributions of clinical characteristics, with the real-world cohorts being more heterogeneous. Our results demonstrate the feasibility of estimating survival in real-world target populations that are substantially more diverse than those enrolled in clinical trials. By statistically adjusting for differences in measured risk factors and structural positivity violations, we show how trial data could potentially be extrapolated to inform real-world patient outcomes. This approach can enhance understanding of the broader public health impact of therapy approvals and reimbursement decisions, with important implications for equity.

Generalizability plays an important role in decision-making by addressing gaps in local data, particularly in contexts where clinical trial evidence may not fully represent the target population and real-world evidence is used to supplement them (He et al. 2020). As regulatory and health technology assessment bodies increasingly rely on real-world data, methods to adjust for cross-population differences in disease characteristics, treatment practice, and healthcare systems may be important to ensure that evidence is both scientifically valid and locally relevant. Indeed, the National Institute for Health and Care Excellence (NICE) has recently updated their real-world evidence guidance (<u>NICE RWE</u> <u>framework</u>) to include specific recommendations for evaluating external validity, highlighting its importance in bridging the gap between trial efficacy and real-world effectiveness to inform timely and equitable access to innovations. Although our study was to assess the generalizability of a clinical trial, the framework can also be adapted to assess the cross-country transportability of real-world evidence (e.g., Ramagopalan et al. 2022). Furthermore, with rigorous validation, such exploratory approaches hold promise for informing the impact of novel healthcare policies in more equitable and inclusive contexts – even when derived from limited, highly selected populations.

Adjustment for measured variables alone (BASE analysis) substantially improved the alignment of real-world survival estimates with those observed in the RCT relative to no adjustment. Given that most key prognostic factors are routinely captured in practice, our findings suggest that adjusting for differences in these variables can meaningfully reduce baseline risk imbalance across populations. Notably, the BASE model more closely approximated survival in the French and German cohorts than in the US cohort. This was counterintuitive because it is more likely that the US trial Lung-MAP S1400I should generalize better to the US real-world population than to populations in other countries that have different demographics and standards of care compared to the United States – indeed, measured patient characteristics were more closely aligned between Lung-MAP S1400I and the US cohort than with Germany or France. These results suggest that while adjustment for measured variables is necessary, it is not sufficient. Researchers looking to implement it for transporting trial findings to Germany or France may need to consider additional cross-country differences that we may have overlooked.

We next used pre-specified parameters for unmeasured/mismeasured variables, including those underlying structural positivity violations (POS analysis). These parameters, namely the variables’ population prevalence and their (conditional) association with mortality on the hazard ratio scale, were estimated from published papers, and in some cases, based on clinical expert opinion when no quantitative external information was available. Compared to the BASE model, the POS model was able to approximate US real-world survival almost perfectly. Approximation to survival from the CRISP registry in Germany and EVIDENS study from France, both of which included large sample sizes, were generally quite good. In both German datasets, there was an underestimation of the mortality risk initially, which may be due to a misspecification of parameters for the baseline factors in the German population. Except for the German CRISP registry, the POS model overestimated mortality risk in Germany and France after approximately 5-10 months of follow-up; this was also observed in the BASE analysis. This is surprising because RCT patients are thought to be healthier and better monitored for adverse events and progression, and we would have expected the reverse – an underestimation of real-world mortality risk when using the trial data to infer real-world outcomes. It is unclear what factor(s) could have led to the model overestimating the risk for all-cause mortality over time in the German and French populations.

Importantly, the control cohorts in Japan (*) and England (*) behaved as expected in all analyses. The confidence intervals were quite wide for Japan in the POS analysis, which may be because of the uncertainty from highly imbalanced variables such as *EGFR*/*ALK* mutations and prior immuno-/targeted therapy that had a very low prevalence in the trial.

The strengths of this study are in a pre-specified study plan, a rigorous handling of positivity issues and unmeasured/mismeasured variables, and the application of synthesis estimators to address structural positivity violations. We found that the target trial specification was useful in identifying and clearly reporting differences in eligibility criteria between the different populations. All assumptions and parameter inputs were transparently documented and defined a priori, ensuring analytical integrity. We also incorporated controls to contextualize study findings – intentionally selected target populations where good and poor approximation of survival based on the trial was expected. Finally, we demonstrated the use of transportability analysis where only summary-level data for patient characteristics is available in the target population, even when the target population is significantly more diverse than the trial.

This study has a few limitations. We used a synthesis of measured data from Lung-MAP S1400I and external information to build the POS model. For some variables, such as presence of *EGFR*/*ALK* mutations, we could not find published studies in a squamous-only NSCLC population that quantified the association of the presence of these variants with overall survival. Given the qualitative differences in some of these associations between squamous and non-squamous NSCLC (see results from multivariable analysis stratified by histology in Stenehjem et al. 2021), we chose not to use values from NSCLC populations that included non-squamous histology. Instead, our synthesis model relies on the clinical expertise about these parameters within our team, which we prespecified prior to the analysis; we have also reported our rationale for the values (see *Appendix – Study specification: Synthesis parameters*).

However, it is possible that we have incorrectly specified the magnitude and/or direction of one or more of these (conditional) associations (also see caveats in Zivich et al. 2024), and that could have an impact on the results. It is possible to pre-specify and implement sophisticated sensitivity analyses to evaluate the impact of these assumptions on the results in cases where external information or real-world data is lacking (Shi et al. 2024).

We also did not adjust for any unmeasured time-varying risk factors for mortality. It is of course possible to simulate time-varying variables to adjust for them, but we chose not to do it in this study primarily because we did not find evidence of significant differences in any time-varying variables between countries (see *Appendix – Justification for other study design choices*). Another limitation is the use of summary-level data. We provide justification and sensitivity analyses for our analytical choices on the use of summary-level data and copula-based simulation, but we cannot discount their misspecification. Further work is needed on their appropriate use for transportability adjustment.

In conclusion, our findings demonstrate that it is possible, under clearly specified assumptions and with appropriate methodological safeguards, to generalize RCT results to more diverse real-world populations. This has important implications for health equity and public health decision-making, where actionable evidence is often needed for underrepresented or systematically excluded groups. Beyond enabling cross-country transportability assessments and more robust external control arm analyses, transportability methods may offer a practical path to designing more inclusive trials by informing broader eligibility criteria or extrapolating evidence to new indications. While these methods cannot substitute for well-powered RCTs in the target populations of interest, they provide a valuable complement when trials are infeasible or unavailable. Future benchmarking across diverse datasets, therapeutic areas, and geographies will be essential to identify the boundaries of statistical methods for transportability, and to support wider adoption of these methods in regulatory and health policy contexts.

## Supporting information

Appendix - Study specifications

Appendix - Statistical formalism

## Data Availability

This work was performed using data from the NCTN Data Archive of the National Cancer Institute's (NCI's) National Clinical Trials Network (NCTN). Data were originally collected from clinical trial NCT number NCT02785952. Data can be requested online via Project Data Sphere.

## Acknowledgements

We thank Paul Zivich (University of North Carolina, Chapel Hill) for his comments on the study plan and encouragement.

## Disclaimer

This work was performed using data from the NCTN Data Archive of the National Cancer Institute’s (NCI’s) National Clinical Trials Network (NCTN). Data were originally collected from clinical trial NCT number NCT02785952. All analyses and conclusions in this manuscript are the sole responsibility of the authors and do not necessarily reflect the opinions or views of the clinical trial investigators, the NCTN, or the NCI.

## References

Anderson, Timothy S., et al. “Generalizability of clinical trials supporting the 2017 American College of Cardiology/American Heart Association blood pressure guideline.” JAMA Internal Medicine 180.5 (2020): 795–797.

Barlesi, Fabrice, et al. “Effectiveness and safety of nivolumab in the treatment of lung cancer patients in France: preliminary results from the real-world EVIDENS study.” Oncoimmunology 9.1 (2020): 1744898.

Chouaid, Christos, et al. “Effectiveness of Nivolumab in Second-Line and Later in Patients with Advanced Non-Small Cell Lung Cancer in Real-Life Practice in France and Germany: Analysis of the ESME-AMLC and CRISP Cohorts.” Cancers 14.24 (2022): 6148.

Degtiar, Irina, and Sherri Rose. “A review of generalizability and transportability.” Annual Review of Statistics and Its Application 10.1 (2023): 501–524.

de Jonghe, Annemarieke, et al. “Underrepresentation of patients with pre-existing cognitive impairment in pharmaceutical trials on prophylactic or therapeutic treatments for delirium: a systematic review.” Journal of Psychosomatic Research 76.3 (2014): 193–199.

Gettinger, Scott N., et al. “Nivolumab plus ipilimumab vs nivolumab for previously treated patients with stage IV squamous cell lung cancer: the lung-MAP S1400I phase 3 randomized clinical trial.” JAMA oncology 7.9 (2021): 1368–1377.

Gupta, Alind, et al. “Transportability of patient outcomes from a US clinical trial to real-world populations-a case study using Lung-MAP S1400I (NCT02785952).” medRxiv (2024): 2024–05.

Gupta, Alind, et al. “Quantitative Bias Analysis for Single-Arm Trials With External Control Arms.” JAMA network open 8.3 (2025): e252152–e252152.

He, Zhe, et al. “Clinical trial generalizability assessment in the big data era: a review.” Clinical and translational science 13.4 (2020): 675–684.

Hernán, Miguel A., Wei Wang, and David E. Leaf. “Target trial emulation: a framework for causal inference from observational data.” Jama 328.24 (2022): 2446–2447.

Inoue, Kosuke, and William Hsu. “Transportability Analysis—A Tool for Extending Trial Results to a Representative Target Population.” JAMA network open 7.1 (2024): e2346302–e2346302.

Kennedy-Martin, Tessa, et al. “A literature review on the representativeness of randomized controlled trial samples and implications for the external validity of trial results.” Trials 16 (2015): 1–14.

Kim, Edward S., et al. “EGFR tyrosine kinase inhibitors for EGFR mutation-positive non-small-cell lung cancer: outcomes in Asian populations.” Future Oncology 17.18 (2021): 2395–2408.

Levy, Natalie S., et al. “Use of transportability methods for real-world evidence generation: a review of current applications.” Journal of Comparative Effectiveness Research 13.11 (2024): e240064.

Loudon, Kirsty, et al. “The PRECIS-2 tool: Designing trials that are fit for purpose.” BMJ 350 (2015).

Mishra-Kalyani, P. S., et al. “External control arms in oncology: current use and future directions.” Annals of Oncology 33.4 (2022): 376–383.

Morita, Ryo, et al. “Real-world effectiveness and safety of nivolumab in patients with non-small cell lung cancer: A multicenter retrospective observational study in Japan.” Lung Cancer 140 (2020): 8–18.

RemirolJAzócar, Antonio. “Transportability of modellJbased estimands in evidence synthesis.” Statistics in Medicine 43.22 (2024): 4217–4249.

Rogers, James R., et al. “Comparison of clinical characteristics between clinical trial participants and nonparticipants using electronic health record data.” JAMA Network Open 4.4 (2021): e214732–e214732.

Rothwell, Peter M. “External validity of randomised controlled trials: “to whom do the results of this trial apply?”.” The Lancet 365.9453 (2005): 82-93.

Schwartz, Daniel, and Joseph Lellouch. “Explanatory and pragmatic attitudes in therapeutical trials.” Journal of chronic diseases 20.8 (1967): 637–648.

Sebastian, Martin, et al. “Prospective, Noninterventional Study of Nivolumab in Real-world Patients With Locally Advanced or Metastatic Non–small Cell Lung Cancer After Prior Chemotherapy (ENLARGE-Lung).” Journal of Immunotherapy 45.2 (2022): 89–99.

Shi, Xiaoting, et al. “Quantitative bias analysis methods for summary level epidemiologic data in the peer-reviewed literature: a systematic review.” Journal of Clinical Epidemiology (2024): 111507.

Snee, Michael, et al. “Treatment patterns and survival outcomes for patients with non-small cell lung cancer in the UK in the preimmunology era: a REAL-Oncology database analysis from the IO Optimise initiative.” BMJ open 11.9 (2021): e046396.

Stenehjem, David D., et al. “Real-world effectiveness of Nivolumab monotherapy after prior systemic therapy in advanced non–small-cell lung cancer in the United States.” Clinical Lung Cancer 22.1 (2021): e35–e47.

Stuart, Elizabeth A., Catherine P. Bradshaw, and Philip J. Leaf. “Assessing the generalizability of randomized trial results to target populations.” Prevention Science 16 (2015): 475–485.

Van Buuren, Stef, and Karin Groothuis-Oudshoorn. “mice: Multivariate imputation by chained equations in R.” Journal of statistical software 45 (2011): 1–67.

Zhou, Wei, and David C. Christiani. “East meets West: ethnic differences in epidemiology and clinical behaviors of lung cancer between East Asians and Caucasians.” Chinese Journal of Cancer 30.5 (2011): 287.

Zivich, Paul N., et al. “Transportability without positivity: a synthesis of statistical and simulation modeling.” Epidemiology 35.1 (2024): 23–31.

