## Appendix for "Global Transportability of Clinical Trial Outcomes to Real-World Lung Cancer Populations A case Study using Lung-MAP S1400I"

**Analysis overview**

The following is a schematic of the analytical workflow used in this study.

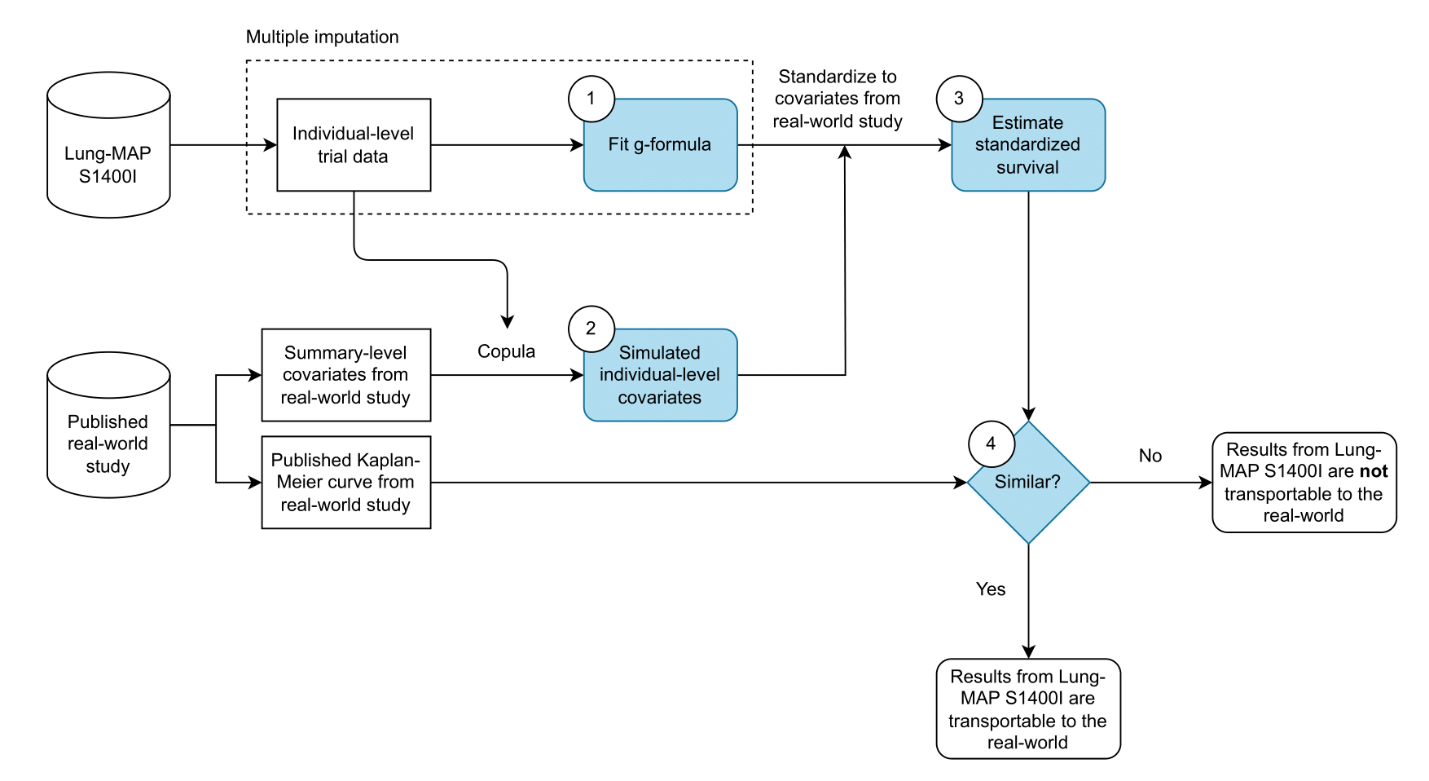

**Identifiability assumptions**

| **Assumption** | **Reasonable for a crude comparison?** | **Analytical plan to address violations** |
| --- | --- | --- |
| Positivity | No | To account for structural positivity violations, a synthesis model using external information will be used. |
| Conditional exchangeability | No | The parametric g-formula will be used to adjust for measured risk factors. Quantitative bias analysis will be performed for unmeasured that do not also underlie positivity violations, if any. |
| No multiple versions of treatment | Yes | There are 2 different versions of nivolumab dosing, as stated in the target trial specification. However, there are no differences in survival or adverse events between these regimens according to [15, 16] and therefore no special analysis is needed as the different treatment versions are not prognostic for the outcome. |
| No interference | Yes | None required |

**Causal graph**

The following causal graph encodes our assumptions about the study setting and was used to guide the choice of variables. All variables in the graph are potentially prognostic for death and would have direct causal edges to the outcome variable “Death”; for simplicity, these edges have not been shown. “Treatment (t0)” refers to treatment assigned or initiated at baseline.

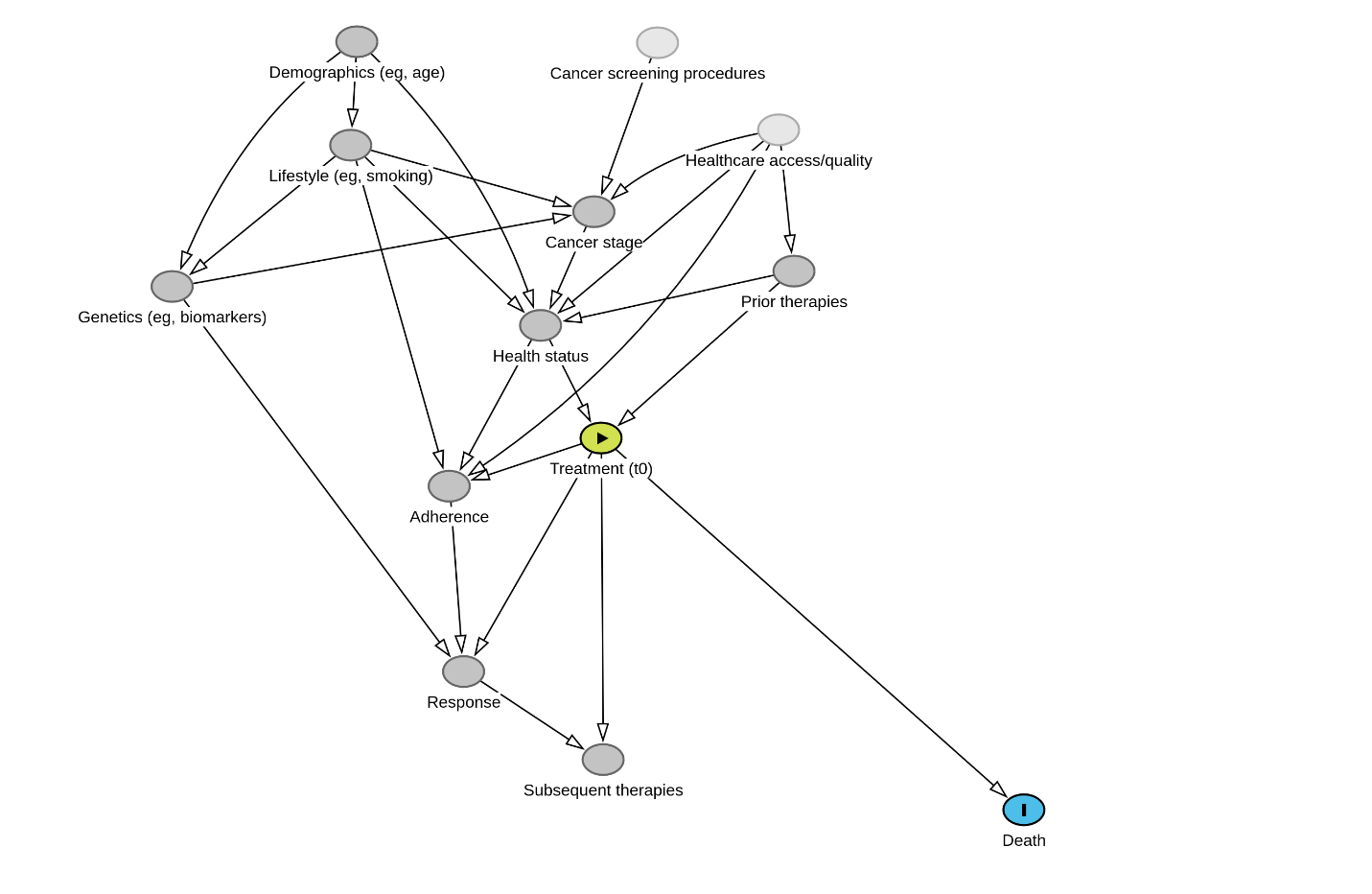

**Study specifications**

Study specification including documentation of target trial elements, data provenance, variables and external parameters can be found in the attached spreadsheet *Transportability study – specificiations.xlsx*.

**Sanity checking**

*Estimation of hazard ratios using standardized survival curves*

As a control, we compared time-averaged hazard ratio estimates using the G-formula to the hazard ratio from the Cox proportional hazards model fit directly to the Lung-MAP S1400I data. Both approaches should theoretically yield similar hazard ratio estimates. We estimated a hazard ratio of 0.89 [0.66, 1.35] for BASE analysis and 0.89 [0.59, 1.31] for POS analysis. Both values were close to the hazard ratio from the unstratified Cox proportional hazard model, which was 0.87 [0.66, 1.16]. Therefore, the approach used in this study provided a reasonable approximation of the hazard ratio from the trial.

*Equivalence between G-formula and Cox proportional hazards coefficients*

The G-formula specification used in our analysis represents an approximation of the Cox proportional hazards model. The following is an example from exploratory analyses comparing coefficients between the two approaches showing that they were similar.

| **Model term (term in bracket represents non-reference category)** | **Cox model** | **G-formula** |
| --- | --- | --- |
| age | -0.276 | -0.307 |
| age:ecog(1) | -0.016 | -0.017 |
| brain(Y) | 0.059 | 0.039 |
| ecog(1) | 1.454 | 1.540 |
| age^2 | 0.002 | 0.002 |
| line(2L) | 0.119 | 0.103 |
| liver(Y) | 0.410 | 0.366 |
| liver(Y):TRT(Nivolumab + Ipilimumab) | -0.012 | 0.092 |
| pdl1(Y) | -0.110 | -0.104 |
| race(White) | 0.036 | 0.006 |
| sex(M) | 0.169 | 0.170 |
| smoker(Ever) | 15.546 | 13.277 |
| TRT(Nivolumab + Ipilimumab) | -0.133 | -0.157 |

*Copula choice*

We used the *VineCopula* R package to compare the Normal, t-, Gumbel, Frank and Clayton copulas for data fit using Bayesian Information Criterion (BIC) (A lower BIC indicates better fit to data). The multivariable Normal copula with a parameter of 0.5 (estimate from *VineCopula*) was chosen for the primary analysis as the standard copula because it is commonly used, and it also happened to have the lowest BIC. As a sensitivity analysis, we repeated all analyses using Clayton copula, which had the highest BIC amongst all copulas tested, and therefore models the covariance structure of the data relatively poorly. The results from this sensitivity analysis visually were virtually identical to the primary analysis reported in this study.

Because we were using the empirical covariance matrix of the Lung-MAP trial to generate pseudo-patient data for real-world populations, there was concern that this covariance structure is not generalizable. Off-diagonal correlations between baseline covariates were quite weak in the Lung-MAP data. We examined data from another publicly available trial dataset (NCT00981058; SQUIRE trial in stage IV squamous NSCLC) with a larger sample size of n=549 and found a similar result for the off-diagonal correlations. Therefore, the covariance structure of the Lung-MAP dataset is probably reasonably generalizable. Regardless, further work is needed to confirm the use of summary-level data for transportability analysis.

*Number of iterations*

The number of replicates used for the analysis was tuned to balance sufficiency and computational time. Results from Lung-MAP S1400I were used as a control to ensure that the overall approach, including the number of iterations used, could recover trial-level survival estimates and showed appropriate coverage of confidence intervals when compared to Kaplan-Meier results. After trial-and-error, 500 iterations of bootstrapping were implemented for the primary analysis, along with 10 replicates of multiple imputation and 10 independent samples from copula. Single imputation resulted in insufficiently reliable coverage of 95% confidence intervals and was therefore not used. The iterations were programmed as follows (pseudocode):

*For each imputed dataset from 1 to 10:*

↳ *For each bootstrap sample from 1 to 500:*

↳ *For each copula sample from 1 to 10:*

↳ *Compute survival estimate*

*Calculate mean and percentile-based 95% confidence intervals*

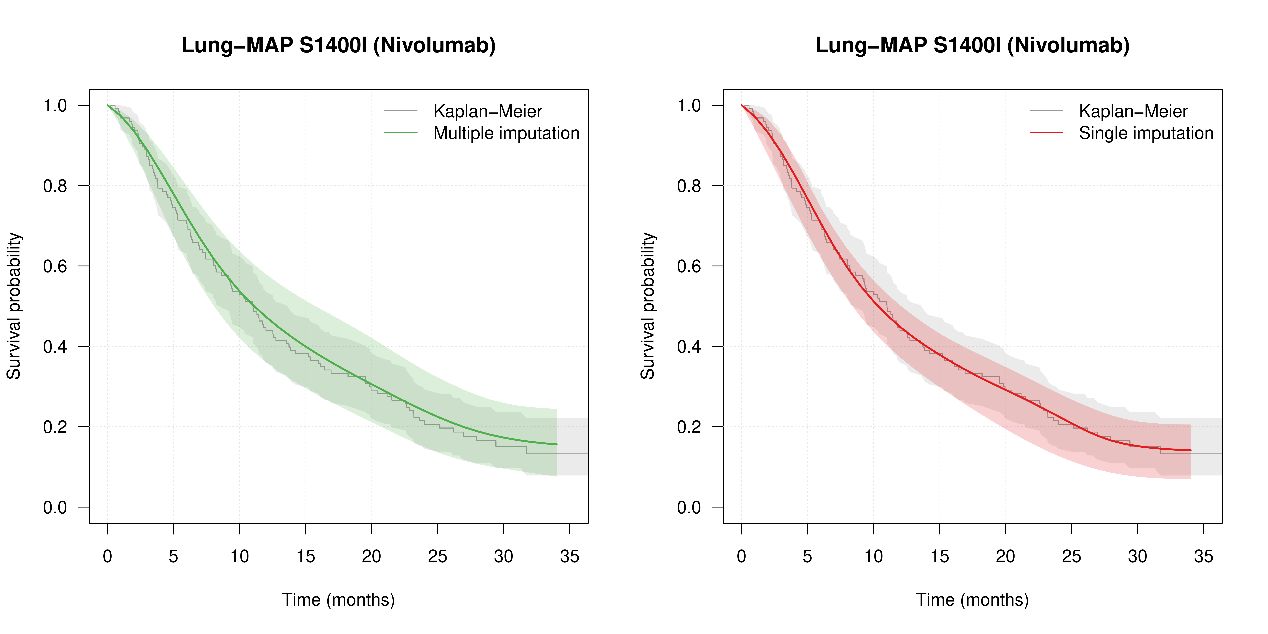

**Justification for other study design choices**

*Differences in treatment adherence*

Because nivolumab is administered intravenously in the inpatient setting and this is an advanced cancer setting, adherence to the prescribed regimen is likely to be relatively high everywhere. Most patients (n= 247 out of 252; 98.0%) in the Lung-MAP study initiated the treatment regimen they were assigned to. Most treatment discontinuations in the trial were due to progression/relapse or adverse events. If we consider non-adherence to include non-initiation of the assigned treatment (n=5), discontinuation for non-protocol reasons (n=9) and refusal to continue medication that was unrelated to progression, relapse or adverse events (n=15), then the non-adherence rate was approximately 11.5%. While it is possible that real-world adherence in the US, France or Germany differs somewhat from Lung-MAP S1400I, it is unlikely that real-world adherence was higher than trial adherence. Without concrete evidence of differences in adherence between countries, we chose to avoid adjusting for it directly, and therefore it was not considered in our analysis. However, if we assume that higher adherence is associated with improved survival, then our model-based survival estimates without having accounted for adherence differences are possibly biased upwards somewhat.

*Selection of patient cohorts representing the target population*

We are assuming in our analysis that the patients who actually received nivolumab in the real-world studies are representative of all patients eligible to receive nivolumab. We think this is a reasonable assumption because nivolumab is approved regardless of PD-L1 expression levels for this indication. (Note that our target population is defined not by the eligibility criteria of Lung-MAP but by the real world setting of patients who are eligible to receive nivolumab.)

Pembrolizumab has been increasingly preferred in the second-line setting, as well as in the front line in combination with platinum-based chemotherapy, particularly for patients with high PD-L1 expression. Therefore, the real-world cohorts of patients who actually received nivolumab may have been selected for patients with low PD-L1 expression. Notably, PD-L1 expression is not associated with overall survival in patients with squamous NSCLC treated with nivolumab [<https://www.nejm.org/doi/full/10.1056/NEJMoa1504627>], and therefore despite selection on treatment, we do not expect a significant impact on results for survival.

Furthermore, pembrolizumab was approved for this indication by the FDA in October 2018. All real-world studies included in this study ended follow-up around 2020, by which time it is likely that the standard of care with regards to the predominance of nivolumab for this indication, was still relatively intact.

*Differences in subsequent therapies*

The Lung-MAP study does not report subsequent therapies and no information on it was available in the NCI dataset. Because differences in subsequent therapy in the Lung-MAP trial and real-world populations globally could potentially affect transportability results, we attempted to quantify whether such differences existed in the first place.

In France (EVIDENS), 38% of the patients who initiated nivolumab subsequently received a different agent, of which gemcitabine (23%), docetaxel (22%) patients and erlotinib (16%) were most common [<https://doi.org/10.1183%2F23120541.00120-2017>]. Those who did receive subsequent therapy were likely selected patients with good performance status to be able to continue systemic therapy, and they had significantly better survival than those who did not in multivariable analysis (Table 4 in the publication, hazard ratio: 2.55, 1.68-3.89). Our prior work on treatment patterns in a large US oncology database also had qualitatively similar results to those reported by EVIDENS in France (unpublished). In Checkmate-017 (second-line nivolumab vs docetaxel in squamous NSCLC; <https://www.nejm.org/doi/full/10.1056/NEJMoa1504627>), which likely reflects a similar standard of care as Lung-MAP S1400I, subsequent therapies received by patients in the nivolumab arm were also largely chemotherapy agents. 36% of patients received a subsequent therapy at the time of publication, a percentage very similar to that reported by EVIDENS in France. Based on these figures, we argue that there are no significant systematic differences in subsequent therapies between trial and real-world populations.

Incidentally, in the Netherlands [<https://www.nature.com/articles/s41598-021-85696-3>], it was found that patients in second-line oncology trials were 1.5 times more likely to have received subsequent therapy compared to those in the real-world. Despite this, crude overall survival between trials and real-world on nivolumab was comparable. This may indicate that subsequent therapy after nivolumab discontinuation is not highly prognostic for survival for this indication.

*Differences in response*

In the Lung-MAP trial, although time-varying data for progression were not available, the best objective response was reported for each patient. 17.5% of patients had complete or partial response, 38.5% had stable disease, and 40.5% showed deterioration as best response, with the remainder being inadequately assessed for it. In the global phase III study Checkmate-017 of nivolumab in squamous cell carcinoma, including patients from USA, France, Germany, United Kingdom as well as other countries, the response rate, which is the number of patients with a complete or partial response, on nivolumab was 20%, which is similar to Lung-MAP S1400I. Note that the patient population in Checkmate-017 includes healthier patients, for example, those who have non-metastatic cancer.

In real-world Korean patients with advanced/metastatic NSCLC, the best response rate was 25% on nivolumab in second line [doi: 10.4143/crt.2020.245], which is higher than the trial. Similarly, in Galician patients, the response rate was reported to be 27.1%, although it included both squamous and non-squamous histologies [doi: 10.21037/tlcr.2018.04.03]. In Japanese patients, it was 20% [DOI: 10.15761/CRR.1000164]. It is unclear what response rates in real-world populations of patients with squamous cell NSCLC who initiate nivolumab were in the populations used in this study, but we assume that any differences would be small (+/-5%). While it is difficult to account directly for differences in response rates because it is a time-varying factor for which little information is directly available for our populations of interest, we do adjust for PD-L1, which is predictive of response to immunotherapy in NSCLC, and based on estimation of tumor mutational burden, which is also predictive of response, which is itself based on the reported or calculated distribution of race/ethnicity, there were no major population differences.

**Changes from protocol version 1.1.1**

1. The ENLARGE-Lung study from Germany was not referenced in the original protocol. It was later identified as a relevant study that had initially been overlooked. As a result, it was subsequently included in the analysis.
2. A preliminary list of covariates for adjustment was included in the protocol. This list was later revised based on a causal graph developed during the analysis phase to better reflect important risk factors.
3. The simple Q-model outlined in the protocol was ultimately not used, as it was deemed overly parsimonious and at risk of model misspecification. A more flexible model that included interactions and higher-order terms was adopted instead.
4. For benchmarking, the “Mean survival probability at the end of follow-up (40 months)” was not computed as a standalone estimate, although it can be visually inferred from the risk curves. Estimates at the end of follow-up were considered imprecise due to limited data at later time points.
5. Similarly, the protocol-specified assessment of “clinically significant differences based on evaluation by oncologist collaborators” was not implemented. It is challenging to judge the clinical significance of differences in survival curves without context. Instead, we interpreted the results considering oncology guidelines on what constitutes a clinically meaningful improvement in median overall survival in lung cancer (see <https://pmc.ncbi.nlm.nih.gov/articles/PMC5632946/> for a mention of suggestion from the American Society of Clinical Oncology (ASCO) for the same).
6. The proposed sensitivity analysis for “Outcome model specification” was not performed. Its stated purpose – to assess sensitivity to unmeasured risk factors – was considered conceptually flawed. Instead, we conducted validation of the model’s estimated parameters to ensure their plausibility and alignment with subject-matter expertise (see Appendix – Sanity checking).
