## Supplementary tables and figures for "Global Transportability of Clinical Trial Outcomes to Real-World Lung Cancer Populations A case Study using Lung-MAP S1400I"

**Supplementary table 1.** Median survival time. The mean and 95% confidence intervals are shown.

| **Target population** | **Follow-up time (months)** | **Median survival time (months)** | | |
| --- | --- | --- | --- | --- |
|  |  | **Actual** | **BASE** | **POS** |
| Lung-MAP S14001 | 35 | 11 [8.6, 3.7] | 10.82 [8.14, 14.00] | 10.54 [7.88, 13.56] |
| USA (Flatiron) | 35 | 7.4 [6.8, 8.5] | 10.40 [8.34, 12.53] | 8.86 [7.10, 11.53] |
| Germany (ENLARGE-Lung) | 23 | 8.9 [6.4, 11.3] | 9.68 [7.07, 13.10] | 9.27 [6.95, 12.43] |
| Germany (CRISP) | 35 | 7.9 [6.1, 11.6] | 9.59 [7.50, 12.37] | 8.85 [6.86, 11.41] |
| France (ESME-AMLC) | 35 | 10.5 [9.6, 12.5] | 10.03 [7.80, 13.24] | 8.95 [6.91, 11.64] |
| France (EVIDENS) | 23 | 10.2 [8.6, 12.1] | 10.09 [8.04, 12.85] | 9.34 [7.43, 12.31] |
| Japan* | 22 | 14.6 [12.3, 15.9] † | 9.42 [6.83, 12.60] | 8.84 [5.00, 14.27] |
| England* | 35 | 4.80 [2.4, 11.9] ‡ | 10.33 [8.33, 12.59] | 8.82 [6.54, 12.04] |

† The number shown here is the median overall survival reported by Morita et al. (2020) in the target population as defined in the study specification which includes both squamous and non-squamous cases. They do provide median survival by histology: for squamous cell NSCLC it was 12.3 [10.5, 14.2] months. However, because they did not provide baseline characteristics stratified by histology, we could not use it for the primary analysis.

‡ Stage IIIB-IV SQ 2013-2017

**Supplementary table 2.** Hazard ratios for all-cause mortality estimated in the POS analysis.

| **Target population** | **Hazard ratio [95% CI]** |
| --- | --- |
| Lung-MAP S14001 | 0.89 [0.59, 1.31] † |
| USA (Flatiron) | 0.86 [0.60, 1.37] |
| Germany (ENLARGE-Lung) | 0.84 [0.47, 1.53] |
| Germany (CRISP) | 0.90 [0.59, 1.47] |
| France (ESME-AMLC) | 0.78 [0.53, 1.43] |
| France (EVIDENS) | 0.75 [0.54, 1.18] |

† The reported hazard ratio in the Lung-MAP trial was 0.87; 95% CI 0.66-1.16 (Gettinger et al. 2021)

**Supplementary figures**

**Supplementary figure 1.** Results from the BASE analysis after adjusting for measured variables only including England*. The grey curve represents crude survival in the Lung-MAP trial, which is the same across all panels. Shaded blue regions represent 95% confidence intervals. Model-based estimates (blue) are plotted along with the observed survival in the real-world cohorts (red).

For England*, the difference in predicted versus actual RMST was 4.48 [2.91, 6.30] months for the BASE analysis, compared to -4.91 months for CRUDE.

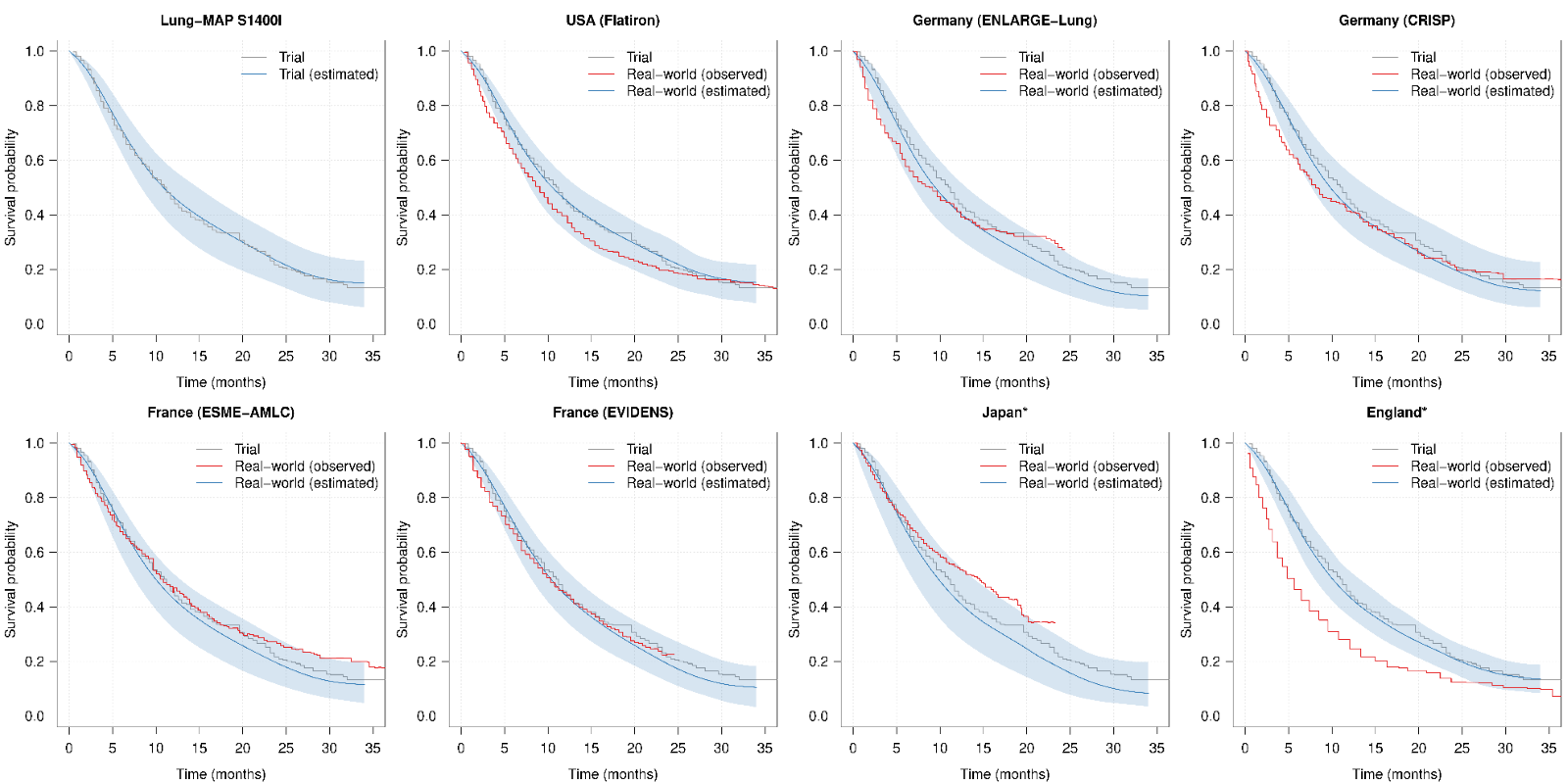

**Supplementary figure 2.** Results from the POS analysis after accounting for positivity violations, including England*. The grey curve represents crude survival in the Lung-MAP trial, which is the same across all panels. Shaded blue regions represent 95% confidence intervals. Model-based estimates (blue) are plotted along with the observed survival in the real-world cohorts (red); a high degree of overlap between the blue and red curves reflects high transportability.

For England*, the difference in predicted versus actual RMST was 3.02 [0.46, 5.52] months for the BASE analysis, compared to -4.91 months for CRUDE.

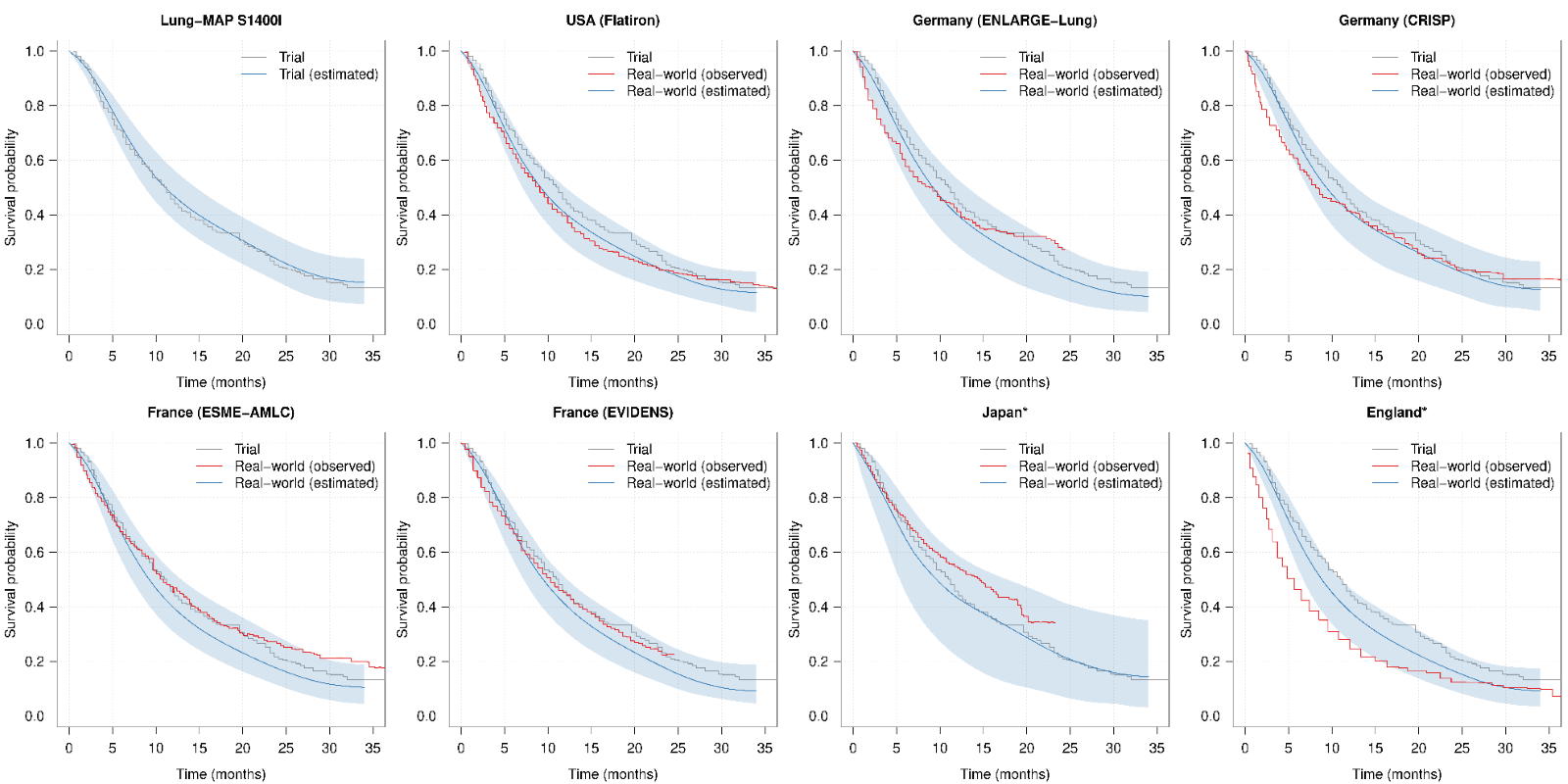
