## Appendix - Statistical formalism for "Global Transportability of Clinical Trial Outcomes to Real-World Lung Cancer Populations A case Study using Lung-MAP S1400I"

### Generalizability of Clinical Trial Outcomes to Global Real-world Populations in Patients with Squamous Cell Lung Cancer

Gupta et al.

Appendix – Statistical formalism

#### 1 Problem setup

- $Y \in \{0, 1\}$ represents the outcome all-cause mortality by (without loss of generality) the end of follow-up where $Y = 1$ represents death.
- $A \in \{0, 1\}$ represents treatment, where $A = 0$ represents initiation of nivolumab monotherapy.
- $P[Y^{a=0} = 1]$ represents the probability of death by the end of follow-up in the population if everyone in the population initiated nivolumab monotherapy.

Our *study population* $S$ consists of the set of patients in the United States who met the eligibility criteria of the Lung-MAP S1400I trial. Patients in the *study sample*, that is those included in the Lung-MAP study, were sampled from this study population $S$. We assume that the study sample is a random subset of the study population. Because $P_S[Y^{a=0} = 1]$ is unobservable, we assume that $\hat{P}_S[Y^{a=0} = 1]$, which is the sample-level distribution of death in Lung-MAP S1400I approximates $P_S[Y^{a=0} = 1]$. Because Lung-MAP S1400I is a randomized trial, we can reframe the counterfactual distribution in terms of observables: $\hat{P}_S[Y^{a=0} = 1] = P_S[Y = 1|A = 0]$, which is simply survival in the trial within the nivolumab arm. We assume $P_S[Y = 1|A = 0]$ is an unbiased estimate of the population-level mortality on nivolumab in $S$.

- Note that we do not invoke the study population further because the goal of the study is to transport trial-level outcomes $P_S[Y = 1|A]$. It is essential to note however the distinction between the study population and our target population of US patients eligible to receive nivolumab based on the Stenehjem et al. (2021) paper. The latter is not equal to nor a subset of the study population. Therefore, even for the US, we are actually interested in transportability, not generalizability.
- Complete data on patient outcomes was measured in the Lung-MAP S1400I study, that is, there were no study dropouts.

We are interested in population-level mortality $P_T[Y^{a=0} = 1]$ in a *target population* which we denoted by $T$. In our study, we include multiple target populations, which are all essentially the set of patients in the United States, Germany and France who are eligible to receive $A = 0$. The real-world studies included in our study sampled (we assume randomly) patients from the target populations in their respective countries, and therefore we have $P_T[Y = 1|A = 0]$ – the observed survival in real-world studies – as estimates for $P_T[Y^{a=0} = 1]$ under the assumption that patients who received nivolumab were representative of the entire real-world population of patients eligible to receive nivolumab (see *Appendix - Selection of patient cohorts representing the target population* for justification).

- We now simply use "real-world target population" to mean both the target population and the sample from the target population, the latter as the observed estimate of the unobserved former.
- Study discontinuation occurs in the real-world studies. For simplicity we assume that censoring $C = 1$ was uninformative. Therefore, even though what we actually measure is $P_T[Y = 1|A = 0, C = 1]$, that it is still an unbiased estimate of $P_T[Y^{a=0} = 1]$.
- Formally, each real-world study may have had slightly different eligibility criteria. For example, within France, the ESME-AMLC cohort excluded patients with concomitant cancers, whereas the EVIDENS cohort did not report such an exclusion. The target populations are therefore defined by the eligibility criteria of the respective real-world study.

#### 2 Goal of transportability analysis

When a clinical trial is performed, we have access to data from the clinical trial, including $P_T[Y = 1|A]$, and may then wish to extrapolate this trial-level outcomes to a real-world target population that is more diverse or inclusive than the trial. $P_T[Y = 1|A]$ is typically not available due to lack of data on patient outcomes $Y$ and/or $A$ in the target population. Therefore, the goal is use the trial data to estimate $P_T[Y = 1|A]$ using trial data.

In our empirical validation study, $P_T[Y = 1|A = 0]$ is available and acts as our gold-standard for benchmarking results from transportability analysis against. That is, we use transportability analysis to try to estimate $P_{S\to T}[Y = 1|A = 0]$ and compare it to $P_T[Y = 1|A = 0]$. If they are similar, we can assert that it is possible to transport results from the trial to the target real-world population.

- As an exploratory analysis, if we find $P_{S\to T}[Y = 1|A = 0] \approx P_T[Y = 1|A = 0]$, then we can also compute $P_{S\to T}[Y = 1|A = 1]$ and calculate a hazard ratio over the follow-up comparing standardized survival on $A = 1$ versus $A = 0$ in the target real-world populations.

How we estimate $P_{S\to T}[Y = 1|A = 0]$ is described in the next section.

#### 3 Transportability analysis using G-computation

- $\boldsymbol{X}$ represents a set of pre-treatment baseline risk factors for the outcome required for exchangeability between the trial and real-world populations. That is, within strata defined by $\boldsymbol{X}$, the mortality risk $P_S[Y^a = 1|\boldsymbol{X}] = P_T[Y^a = 1|\boldsymbol{X}]$ is constant across the study and target populations. (See *Appendix* sections *Differences in subsequent therapies* and *Differences in response* for why we ignored time-varying risk factors)
- $P_S[\boldsymbol{X} = \boldsymbol{x}]$ is the observed distribution of these variables in the study sample and $P_T[\boldsymbol{X} = \boldsymbol{x}]$ in the target population, again assuming that the sample distributions are representative to the (unobserved) population-level distributions. Notably, $\hat{P}_T[\boldsymbol{X} = \boldsymbol{x}]$ is often available even if data on treatment and outcomes in the target real-world population are not.

As one can imagine, the number of risk factors $X \in \boldsymbol{X}$ required to fulfill this requirement may be so large as to render the approach for adjusting for them to be a non-starter. For practicality, we assume that only the most important risk factors for mortality that differ in their distributions between the study and target populations are necessary for adjustment.

#### 3.1 G-computation when positivity holds

We assume consistency of outcomes, good measurement, and, for now, that positivity holds. That is, $P_S[\boldsymbol{X} = \boldsymbol{x}] > 0$ for any strata defined by $X \in \boldsymbol{X}$ for which $P_T[\boldsymbol{X} = \boldsymbol{x}] > 0$. Simply put, we must have measured data for all covariate strata defined by $\boldsymbol{X}$ in the trial for which we wish to estimate outcomes for in the real-world population.

The non-parametric version of the G-formula uses standardization to estimate the population-level outcomes. That is,

$$P_T[Y = 1|A] = \sum_{\boldsymbol{x}} P_T[Y = 1|A, \boldsymbol{X} = \boldsymbol{x}] P_T[\boldsymbol{X} = \boldsymbol{x}]$$

Essentially we are standardizing the strata-level risks by the prevalence of each stratum in the population.

By our definition of conditional exchangeability,

$$P_T[Y = 1|A = 0, \boldsymbol{X}] = P_S[Y = 1|A = 0, \boldsymbol{X}]$$

Therefore, we can estimate it using trial data. Because non-parametric estimation can be challenging with the presence of continuous-valued variables, we use the parametric vetheyrsion of the G-formula and estimate $P[Y|A, \boldsymbol{X}]$ using, in our case, a logistic regression model. That is,

$$\hat{P}_S[Y = 1|A = 0, \boldsymbol{X}] = \frac{1}{1 + \exp(-\hat{\boldsymbol{\beta}}^t \boldsymbol{X})}$$

- The superscript $^t$ here represents the transpose.
- The regression model specification used in our primary analysis includes additionally, interactions and higher-order terms.
- The specification for a pooled logistic regression model additionally includes time as a covariate.

Here, $\hat{\boldsymbol{\beta}}$ are the parameters of this model are estimated from the trial data using pooled logistic regression. The model can then be used to predict the standardized survival probabilities in the target real-world population based on the observed distribution of covariates in the target population $\boldsymbol{X} = \boldsymbol{x}_i \forall i \in N$ and then taking their average.

- Due to lack of patient-level data in the target population, we sample this patient-level distribution of risk factors $\boldsymbol{x}_i \sim \hat{P}_T[\boldsymbol{X}]; \boldsymbol{\theta}_{cop}$ from a copula as described in the main text with copula parameters $\boldsymbol{\theta}_{cop}$ estimated from trial data.

This gives us $\hat{P}_{S \to T}[Y = 1|A = 0]$ parametrized by $\hat{\boldsymbol{\beta}}$. We can finally compare this to $P_T[Y = 1|A = 0]$ to assess whether transportability has been sufficiently "achieved". 95% confidence intervals can be estimated by bootstrapping.

#### 3.2 Synthesis estimators for lack of positivity

- Assume now that $\boldsymbol{X} = \{\boldsymbol{U}, \boldsymbol{V}\}$ (all dichotomous variables) where $\boldsymbol{U}$ is the set of variables for which all levels $U = \{0, 1\} \forall U \in \boldsymbol{U}$ that are observed in the real-world target population are also observed in the study sample. On the other hand, $\boldsymbol{V}$ is the set of variables for which, let's assume, that only $V = 0 \forall V \in \boldsymbol{V}$ is observed in the trial data, whereas $V = \{0, 1\} \forall U \in \boldsymbol{U}$ are represented in the real-world target population. Clearly then, $P_S[\boldsymbol{U}, \boldsymbol{V} = 1] = 0$ even though $P_T[\boldsymbol{U}, \boldsymbol{V} = 1] > 0$.
  - For example, one such variable $V$ is ECOG performance score, for which an ECOG score of 0 or 1 is observed in the trial ($V = 0$) but a score of 2+ ($V = 1$) is not because it is an exclusion criterion. The probability of observing patients with an ECOG score of 2 or higher in Lung-MAP S1400I is zero by definition.

Given this limitation, it is not possible to estimate $\hat{P}_{S \to T}[Y = 1|A = 0, \boldsymbol{X}] \equiv \hat{P}_{S \to T}[Y = 1|A = 0, \boldsymbol{U}, \boldsymbol{V}]$ but only $\hat{P}_{S \to T}[Y = 1|A = 0, \boldsymbol{U}, \boldsymbol{V} = 0]$.

The synthesis estimator for addressing positivity violations described in Zivich et al. (2024) works by updating, in our case, the predictions from the logistic regression model estimated from trial data conditional on $\boldsymbol{V} = 0$ with parameters $\hat{\boldsymbol{\beta}}$ with an additional set of parameters $\hat{\boldsymbol{\alpha}}$ that need to be estimated using external information such that:

$$\hat{P}_S[Y = 1|A = 0, \boldsymbol{X}] = \hat{P}_S[Y = 1|A = 0, \boldsymbol{U}, \boldsymbol{V}] = \frac{1}{1 + \exp(-\hat{\boldsymbol{\beta}}^t \boldsymbol{U} - \hat{\boldsymbol{\alpha}}^t \boldsymbol{V})}$$

In our case,for example, the value of $\hat{\alpha}$ for ECOG performance score then can be approximated by the coefficient for that variable from a multivariable Cox model in a similar population that includes patients with both ECOG score categories of 0-1 (reference) and 2+. The rationale for our choices of $\hat{\boldsymbol{\alpha}}$ can be found in *Appendix - Study specifications*. For details on synthesis estimators, see Zivich et al. (2024).

### 4 Controls

A *control* refers to a target population $T'$ receiving treatment $A'$ in which it is expected, based on subject-matter knowledge, that $P_{S \to T'}[Y = 1|A = a]$ will closely approximate $P_{T'}[Y^{a'} = 1]$ (positive control) or not (negative control) under the exact set of assumptions and model specification as in the primary analysis. This may be the case, for example, because we know that either all identifiability and estimatability conditions are met (in the case of a positive control $\{T', A'\}$) or because one or more identifiability conditions are not met (in the case of a negative control $\{T', A'\}$). Here, treatments (or treatment regimens) $A'$ may be different from $A$.

#### 4.1 Positive control

Assume that the target population $T'$ is the same as the study population $S$. Naturally, one expects that $P_{S \to S}[Y = 1|A = a]$ should closely approximate $P_S[Y = 1|A = a]$ given the same transportability model as the primary analysis.

- Note that in fact all identifiability assumptions are trivially satisfied when transporting from $S \to S$ and indeed the set of variables required for conditional exchangeability is actually $\boldsymbol{X} = \emptyset$. However, as we noted in our definition of a control, a control must act as a control under the same model as the primary analysis.

Therefore, if our transportability model is an unbiased estimator of $P_S[Y=1|A=0,\boldsymbol{X}]$, we expect that $\hat{P}_{S\to S}[Y=1|A=a];\hat{\boldsymbol{\beta}} \approx P_S[Y=1|A=a]$ (for the POS analysis, naturally ; $\hat{\boldsymbol{\beta}},\hat{\boldsymbol{\alpha}}$).

- Note that this is necessary but not sufficient for a well-specified transportability model. For example, a model that includes only an intercept term will suffice because $\boldsymbol{X}=\emptyset$ when $T=S$, but in general it will be necessary to adjust for many risk factors for a well-specified transportability model when $T \neq S$, as is the case for the primary analysis in our study.

#### 4.2 Negative control - Morita et al. (Japan)

For the Japanese target real-world population, note that although the study population includes only patients with squamous cell lung cancer, the target population includes, in addition to other more diverse patients common to the primary analysis, patients with non-squamous histology which is the predominant lung cancer subtype accounting for the majority of diagnosed cases. Here, we anticipate that the Japanese population acts as a negative control where $P_{S\to T'}[Y=1|A=0];\hat{\boldsymbol{\beta}} < P_{T'}[Y=1|A=0]$ because:

1. The primary analysis transportability model does not account for positivity violation due to histology in populations in the United States, Germany or France.
2. Histological type is an important risk factor for mortality in patients with lung cancer. Non-squamous histology is associated with longer survival compared to squamous histology.
3. Therefore, the Japanese population includes patients with non-squamous histology who are expected to have better outcomes, on average, than those in the study population — above and beyond what is captured by the transportability model in the primary analysis, which, by design, does not account for histology.

The reason for the Japanese population being a good negative control is therefore because an identifiability condition, that of positivity, is not met and it not being ignorable.

#### 4.3 Negative control - Snee et al. (England) [Appendix-only]

In our definition of a control, and within the transportability framework, it is theoretically possible to conjecture about the transportability of an outcome $Y$ from any study population on a treatment regimen $A$ to any target population on a treatment regimen $A'$. A negative control is simply $\{T',A'\}$ for which one or more identifiability or estimatability conditions do not hold.

Our rationale for including the English study as a negative control in the study protocol was therefore straightforward – it included different eligibility criteria and treatment regimen than those in the study population. The eligibility criteria are detailed in the target trial specification. The treatment regimen $A'$ is simply "Follow the standard of care" instead of $A=0$ which is "Initiate nivolumab monotherapy" while $\hat{P}_{S\to T'}[Y=1|A=0];\hat{\boldsymbol{\beta}}$ where $T'$ =England* estimates the hypothetical outcomes measured for the time of diagnosis in these patients if we could administer nivolumab monotherapy to all eligible patients in $T'$. Importantly, the outcome $Y$ is still measured in the same way in this population as in the study population, that is, from the time of meeting eligibility criteria over a number of months over the follow-up. Clearly, neither positivity nor consistency of outcomes holds for this target population, and therefore this population could theoretically be used as a negative control where the expectation is that $\hat{P}_{S\to T'}[Y=1|A=0];\hat{\boldsymbol{\beta}} > P_{T'}[Y=1|A=0]$. This is because 37.5% of these patients in Snee et al. (2021) were possibly too frail to initiate systemic therapy and therefore the population at diagnosis on average is likely substantially more frail than the patients robust enough to have survived until second-line or later in the study population who were still eligible to continue systemic therapy.

However, due to differences in the timing of eligibility – occurring after progression on one or more lines of therapy in the study population versus at initial diagnosis in this target population – it is not meaningful to adjust for line of therapy, a variable included in the primary analysis model. Since our definition of a control requires using the same transportability model specification as the primary analysis, this population is technically inadmissible as a negative control. Therefore, we excluded results for this cohort from the main text, but provide them in the Appendix for reference.
